# Insights into the comorbidity between type 2 diabetes and osteoarthritis

**DOI:** 10.1101/2023.03.28.23287861

**Authors:** Ana Luiza de S. V. Arruda, April Hartley, Georgia Katsoula, George Davey Smith, Andrew P. Morris, Eleftheria Zeggini

**Affiliations:** Institute of Translational Genomics, Helmholtz Munich, Neuherberg, 85764, Germany; Munich School for Data Science, Helmholtz Munich, Neuherberg, 85764, Germany; TUM school of medicine, Technical University Munich and Klinikum Rechts der Isar, Munich, 81675, Germany; Technical University of Munich (TUM), School of Medicine, Graduate School of Experimental Medicine, Munich, 81675, Germany; MRC Integrative Epidemiology Unit, University of Bristol, Bristol, BS8 2BN, United Kingdom; Centre for Genetics and Genomics Versus Arthritis, Centre for Musculoskeletal Research, The University of Manchester, Manchester, M13 9PT, United Kingdom

## Abstract

Multimorbidity is a rising public health challenge with important implications for health management and policy. The most common multimorbidity pattern is for the combination of cardiometabolic and osteoarticular diseases. Here, we study the genetic underpinning of the comorbidity between type 2 diabetes and osteoarthritis. We find genome-wide genetic correlation between the two diseases, and robust evidence for association signal colocalization at 18 genomic regions. We integrate multi-omics and functional information to resolve the colocalizing signals, and identify high-confidence effector genes, including *FTO* and *IRX3*, which provide proof-of-concept insights into the epidemiologic link between obesity and both diseases. We find enrichment for lipid metabolism and skeletal formation pathways for signals underpinning the knee and hip osteoarthritis comorbidities with type 2 diabetes, respectively. Causal inference analysis identifies complex effects of tissue-specific gene expression on comorbidity outcomes. Our findings provide insights into the biological basis for the type 2 diabetes-osteoarthritis disease co-occurrence.

## Introduction

Multimorbidity is defined as the coexistence of multiple chronic diseases in a single individual [1]. Worldwide, over 50% of the population older than 65 years is affected by more than one long-term medical condition simultaneously [2]. Commensurate with the rise in life expectancy and average population age, multimorbidity is an increasing global health challenge. However, the majority of health and drug development research is focused on treating and/or preventing individual diseases, leading to interventions that are currently not optimally designed to assist patients suffering from multiple health conditions.

The most prevalent multimorbidity pattern among women and men is the combination of cardiometabolic and osteoarticular diseases [3], exemplified by the highly prevalent co-occurrence of type 2 diabetes and osteoarthritis [4]. Between 2009 and 2016, approximately one in three adults with prediabetes in the US suffered from arthritis [5]. Osteoarthritis is the most common whole-joint chronic disorder, affecting over 520 million people worldwide [6]. It is a degenerative disorder characterized by a local and systemic low-grade inflammation state, irreversible loss of cartilage, and additional bone formation that results in pain, its most prevalent symptom [7]. Across the globe, type 2 diabetes affects over 430 million people and is characterized by elevated blood glucose levels and insulin resistance [6]. Both osteoarthritis and type 2 diabetes are complex diseases influenced by genetic, demographic and lifestyle factors, such as older age and obesity [8].

The majority of observational studies have reported a positive epidemiological association between type 2 diabetes and osteoarthritis of the hip or knee [4]. In a meta-analysis including 1,040,175 patients, the unadjusted odds ratio (OR) for type 2 diabetes in osteoarthritis patients compared to non-osteoarthritis patients was 1.41 (95% confidence interval (CI)=[1.21, 1.65]) [9]. For type 2 diabetes patients, the overall risk of osteoarthritis was also higher compared to individuals without type 2 diabetes (unadjusted OR=1.46, 95%CI=[1.08, 1.96], N=32,137) [9]. Articular joint-specific analyses have shown a stronger link between type 2 diabetes and knee osteoarthritis than hip osteoarthritis [9].

Mendelian randomization analyses [10] suggest no causal relation between liability to type 2 diabetes and knee osteoarthritis [11], whereas body-mass index (BMI) has been shown to be causal for both diseases [12], [13]. When adjusting for BMI, studies linking type 2 diabetes and osteoarthritis have yielded conflicting results [4], [9], [14]. Considering that obesity is a major risk factor for both diseases studied here, genetic variants associated with different physiological characteristics of increased adiposity are expected to be shared risk variants for the comorbidity. However, those variants could exert their effects on the comorbidity through alternative biological pathways to obesity through horizontal pleiotropy [10].

Given the increase of the world’s elderly population and the chronic nature of this highly prevalent pair of diseases, understanding their shared genetic background is important in order to identify risk variants and effector genes that could be used as biomarkers or druggable targets for bilateral treatment. Here, we perform a systematic overlap analysis on a genome-wide scale to disentangle the shared genetic aetiology of the type 2 diabetes-osteoarthritis comorbidity, including integration with functional genomics data in relevant cell types, and provide insights into common underpinning mechanisms of disease development, including but not limited to, adiposity.

## Results

### Insights into disease biology and treatment targets

We first assessed the genetic correlation between type 2 diabetes (*N*_*cases*_ = 74,124, *N*_*controls*_ = 824,006) and osteoarthritis (knee: *N*_*cases*_= 62,497, *N*_*controls*_ = 333,557; hip: *N*_*cases*_= 36,445, *N*_*controls*_ = 316,943) on a genome-wide scale using data from the largest GWAS meta-analyses to date (Table S1 and Figure S1). In line with epidemiological evidence, we find greater magnitude of genetic correlation between type 2 diabetes and knee osteoarthritis (r^2^=0.241, SE=0.028, p=2.65e-18) compared to osteoarthritis of the hip (r^2^=0.078, SE= 0.029, p=0.008) (Figure 1A). To assess the potential for bias due to overlapping samples and different sample sizes, we also performed a permutation-based analysis (empirical p-value for knee=0.005, empirical p-value for hip=0.142) (Figure 1B, Table S1 and Figure S2). Causal inference analyses using Mendelian randomization showed evidence for a non-causal relationship between the two diseases (Table S6), consistent with smaller-scale studies in the literature [12].

**Figure 1:**
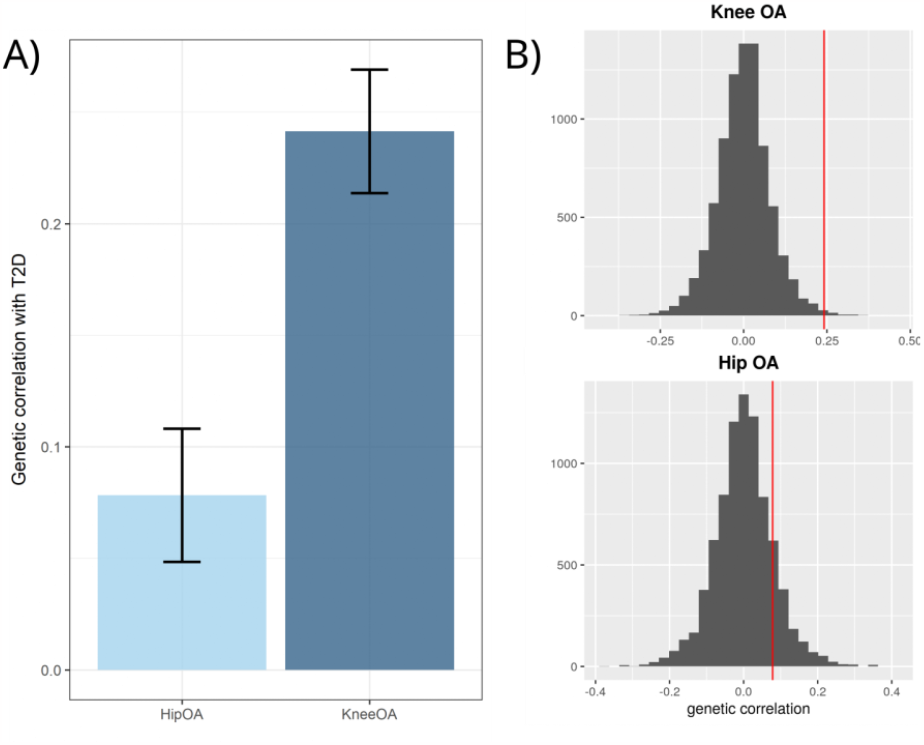
Stronger evidence for a genetic correlation between type 2 diabetes and knee osteoarthritis compared to hip osteoarthritis. A) Genetic correlation results between type 2 diabetes (T2D) and knee or hip osteoarthritis (OA). B) Permutation-based testing results for knee OA and hip OA, respectively. The red line is the true correlation.

Using pairwise Bayesian colocalization analyses on genome-wide significant regions for osteoarthritis (*P* = 1.3*x*10^−8^) or type 2 diabetes (*P* = 5*x*10^−8^), we found robust evidence (posterior probability ≥ 0.8) for a shared causal signal between both diseases at 18 genomic loci (Table S2 and Figures S5-S22). Ten of those loci colocalize with type 2 diabetes for both hip and knee osteoarthritis, two colocalize for hip osteoarthritis only, and six colocalize only for knee osteoarthritis. In three genomic loci, the 95% credible set for the causal variant from the colocalization analysis consisted of a single variant (Figure 2).

**Figure 2:**
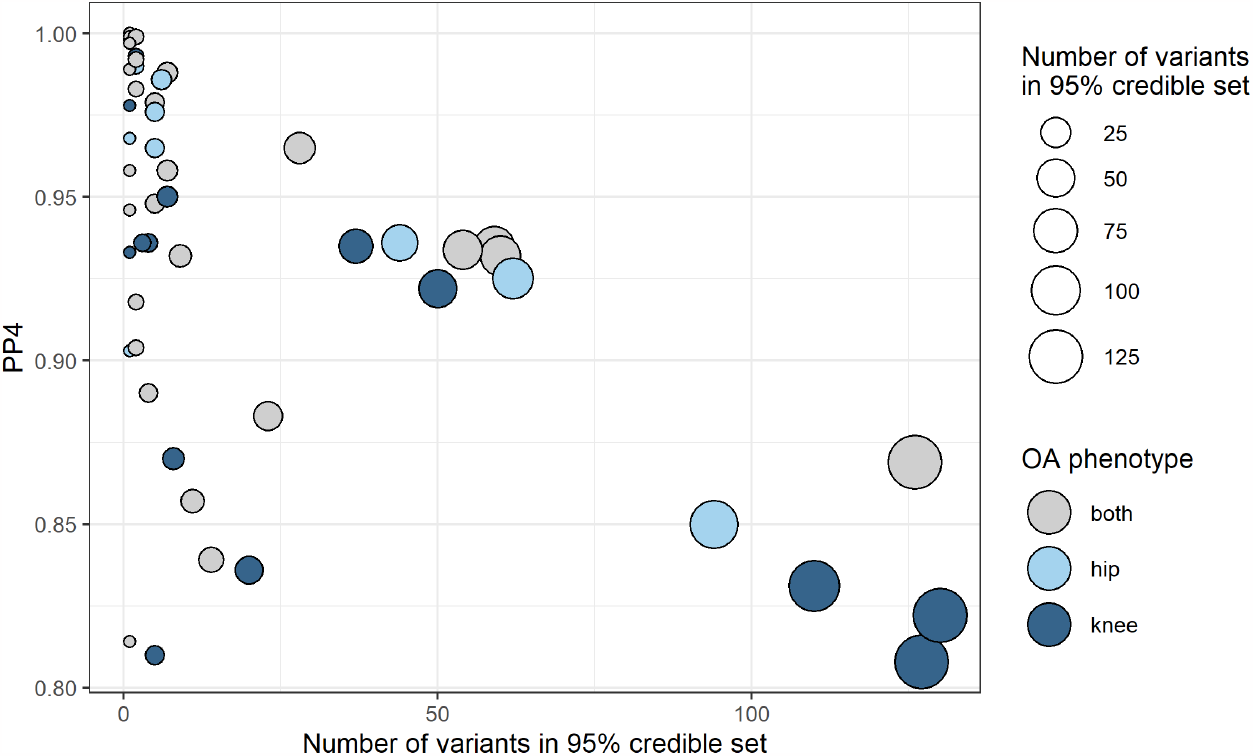
Overview of colocalizing regions. The y-axis depicts the posterior probability of a shared causal variant (PP4) and the x-axis the number of variants in the 95% credible set for the causal variant. Each point represents a colocalized signal between type 2 diabetes and one osteoarthritis (OA) phenotype. Point size is proportional to the number of variants in the colocalization analysis 95% credible set. We find strong statistical evidence for colocalization (PP4>0.8) at 18 unique genomic loci. Some of those loci colocalize between type 2 diabetes and more than one osteoarthrtis phenotype, thus we have 51 points.

We incorporated multi-omics and functional information to identify shared high-confidence candidate effector genes for the type 2 diabetes-osteoarthritis comorbidity in the colocalizing loci. Twelve of these showed statistical evidence for colocalization with gene expression or protein quantitative trait loci (eQTLs and pQTLs, respectively) from disease-relevant tissues (cartilage chondrocytes, synoviocytes and/or pancreatic beta cells). In total, we analysed 906 genes in the vicinity of colocalizing genomic regions by integrating six lines of complementary evidence (Figure 3 and Table S3).

**Figure 3:**
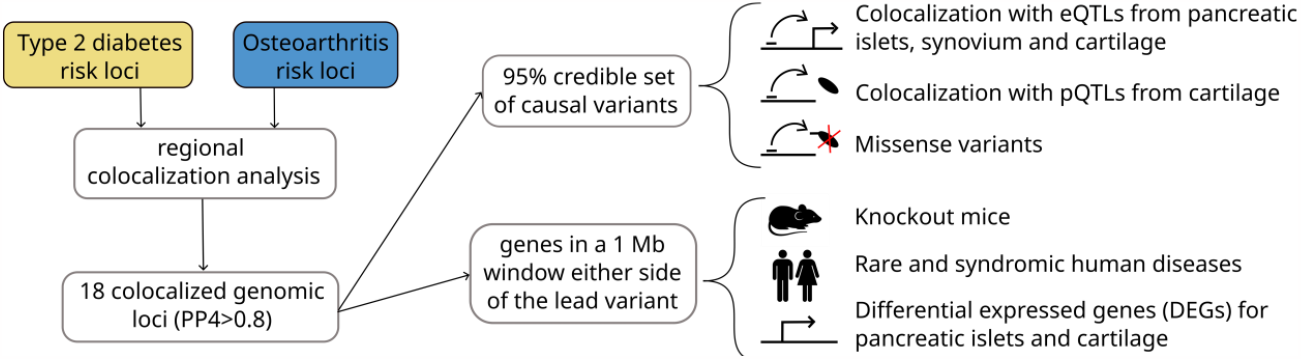
Study design. In each of the 18 genomic loci that colocalized with a posterior probability of a single shared causal variant (PP4) ≥ 0.8, we explored all genes in a 1Mb window on either side of the lead variants of the 95% credible set for the causal variant of the colocalization analysis. For each gene, we searched databases for knockout mice, and rare and syndromic human diseases for pre-defined type 2 diabetes- and musculoskeletal-related phenotypes. We also examined differentially expressed genes (DEGs) in pancreatic islets of healthy versus diabetes patients, and of degraded versus intact osteoarthritis cartilage. We examined the variants in the 95% credible set for the causal variant of each colocalization locus for missense variants. We performed regional multi-trait colocalization analyses between type 2 diabetes, each osteoarthritis phenotype, and molecular QTL (gene expression and protein QTL) from disease-relevant tissues. We also examined previously curated high-confidence effector genes for type 2 diabetes or osteoarthritis (Methods).

We defined 72 genes as likely effector genes for the type 2 diabetes-osteoarthritis comorbidity, as they displayed at least one line of supporting evidence for being involved in both diseases. Of the 72 likely effector genes, 19 showed at least three lines of evidence and were defined as high-confidence effector genes (Figure 4). These represent relevant candidates for further functional and clinical research. Eleven of these have not previously been defined as high-confidence genes for either disease. For two of the high-confidence genes, *APOE* and *WSCD2*, the 95% credible set for the causal variant from the colocalization analysis includes missense variants, namely rs429358 and rs3764002. Six out of 19 high-confidence effector genes are the nearest gene to the lead variant of the respective colocalizing genomic locus: *WSCD2, TCF7L2, JADE2, GLIS3, FTO* and *APOE* (Figure S3).

**Figure 4:**
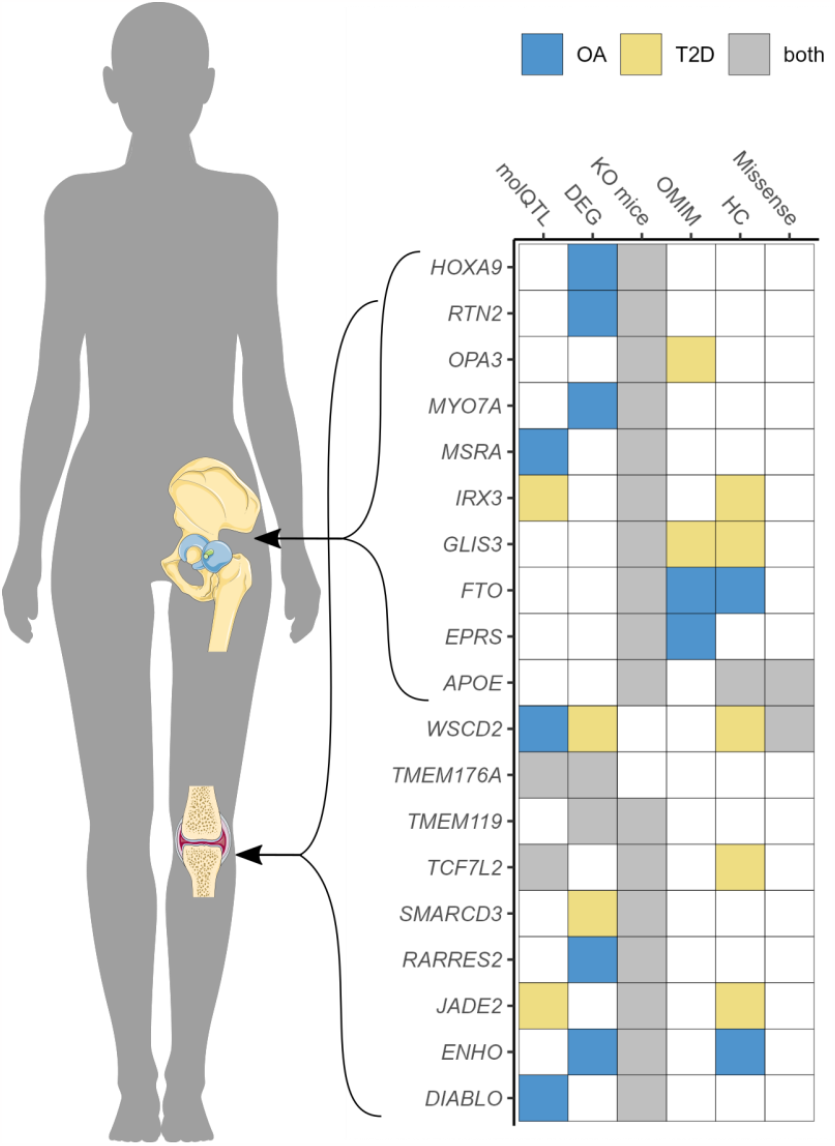
Overview of the 19 high-confidence effector genes for the type 2 diabetes and osteoarthritis comorbidity. Genes are stratified based on the joint affected by osteoarthritis. The scoring of the six biological lines of evidence is depicted on the right (Methods.) (OA = osteoarthritis; T2D = type 2 diabetes; molQTL = molecular quantitative trait loci; DEG = differential expressed genes; KO mice = knockout mice; OMIM = Online Mendelian Inheritance in Man; HC = previously defined high-confidence effector genes; missense = missense variant).

We searched the druggable genome for the druggability status of the likely effector genes for the type 2 diabetes and osteoarthritis comorbidity [15]. Sixteen out of 72 genes were included in the druggable genome (Table S4). Of these, 6 genes are tier 1 druggable targets (*GIPR, TPO, PAK1, SIGMAR1, CTSB, NOS3*), i.e., they are targets of drugs that have market authorization or are in clinical development. The *GIPR* agonist tirzepatide was recently approved for the treatment of type 2 diabetes in adults. It has glucose-lowering effects and has been shown to increase insulin sensitivity [16]. The *PAK1* inhibitor fostamatinib has been approved for the treatment of chronic immune thrombocytopenia [17]. It is also in clinical trial for the treatment of rheumatoid arthritis in order to alleviate the degree of inflammation of the joints [18]. *SIGMAR1* is a target of multiple approved drugs, including pentazocine, which is an analgesic used to treat moderate to severe pain. Naltrexone, an antagonist used in opioid overdose that also targets *SIGMAR1*, is being investigated for treating obesity [19], [20]. The *TPO* gene encodes the thyroid peroxidase protein, which is the target of several approved drugs for the treatment of hyperthyroidism. One of these, the thyroid hormone dextrothyroxine, has been shown to lower serum levels of cholesterol in humans, but the interventional study has been discontinued due to serious adverse effects [21].

The 72 likely effector genes were enriched for several metabolic and cellular process, and for lipid localization and storage pathways. Hip osteoarthritis-related likely effector genes were enriched for bone development pathways and metabolic processes. The 19 high-confidence effector genes were enriched for biological pathways related to diet and obesity (response to caloric restriction and Fto obesity variant mechanism) and for regulation of cell differentiation. The high-confidence genes related to hip osteoarthritis were enriched for the Fto obesity variant mechanism pathway, regulation of lipid localization, and for a biological pathway related to skeletal formation (proximal/distal pattern formation) (Table S5 and Figure S4). These results provide biological support for the link between obesity and both diseases, and for the association between bone development and hip osteoarthritis [7].

### Disentangling the effect of obesity

To explore the role of obesity on the co-occurrence of type 2 diabetes and osteoarthritis, we studied four different measures that capture different aspects of obesity and adiposity: BMI, waist-to-hip ratio, whole body fat mass, and body fat percentage. Sixteen out of the 18 genomic regions that colocalized between type 2 diabetes and osteoarthritis show evidence of association or colocalization (PP4 > 0.8) with at least one adiposity-related trait (Table S9). Four high-confidence effector genes reside in the two genomic regions that do not show any evidence of colocalization or association with the analysed measures of adiposity: *TMEM176A, RARRES2, SMARCD3* and *GLIS3*. These may point to alternative biological mechanisms other than adiposity in the comorbidity between type 2 diabetes and osteoarthritis for these colocalizing signals.

We investigated whether these adiposity measures were causally associated to the expression of high-confidence effector genes in disease-relevant tissues. Within the constraints of the available instruments (Methods), we found evidence of a causal relationship between several adiposity measures and nine high-confidence effector genes (Table S7). For example, we find that all measures of adiposity have a causal effect on higher expression of *IRX3* in synovium or pancreatic islets and on lower expression of *RTN2* in osteoarthritis cartilage. For the high-confidence effector genes located in the two genomic loci that did not show evidence of association or colocalization with adiposity, the direction of effect was not consistent across the different measures employed.

We assessed the causal role of BMI-associated variants with tissue-specific effects, selected based on evidence of their colocalization with brain or subcutaneous adipose tissue eQTLs [22]. For type 2 diabetes, we replicated previous results and showed that BMI-associated variants influencing genes expressed in brain tissue exert a stronger effect on the disease than adipose-tissue related variants [22], although confidence intervals largely overlapped. For knee osteoarthritis, we observed the same trend (Figure S23). For hip osteoarthritis, the results of the causal inference analysis provide evidence for a stronger effect of BMI-associated variants that colocalize with adipose-tissue eQTLs than with brain eQTLs (Table S10). Our results suggest a similar biological underpinning of the adiposity effect captured by BMI on type 2 diabetes and knee osteoarthritis, but potentially different processes for hip osteoarthritis.

### Insights gained from individual loci

#### FTO and IRX3

The obesity-related *FTO* locus colocalizes for type 2 diabetes and osteoarthritis with a posterior probability of a shared causal variant of over 92% (Figure 5). The 95% credible set from the colocalization analysis consists of multiple variants in high linkage disequilibrium with each other. The risk-increasing alleles for the lead causal variants are the same across type 2 diabetes and osteoarthritis. In addition to *FTO*, this locus is associated with a further high-confidence effector gene, *IRX3. IRX3* eQTLs in pancreatic islets colocalize with type 2 diabetes and osteoarthritis genetic signals with a PP4 > 0.8.

As shown above, adiposity is causally associated with an increase in *IRX3* expression in pancreatic islets and synovium (Table S7A). Here, we performed causal inference analyses between the expression of high-confidence genes at this locus and type 2 diabetes or osteoarthritis. We find evidence for a causal effect of increased expression of *IRX3* in pancreatic islets on increased risk of type 2 diabetes (OR=1.16; 95%CI=[1.08, 1.25]; p-value=4.4E-05, F-stat= 16.7).

**Figure 5:**
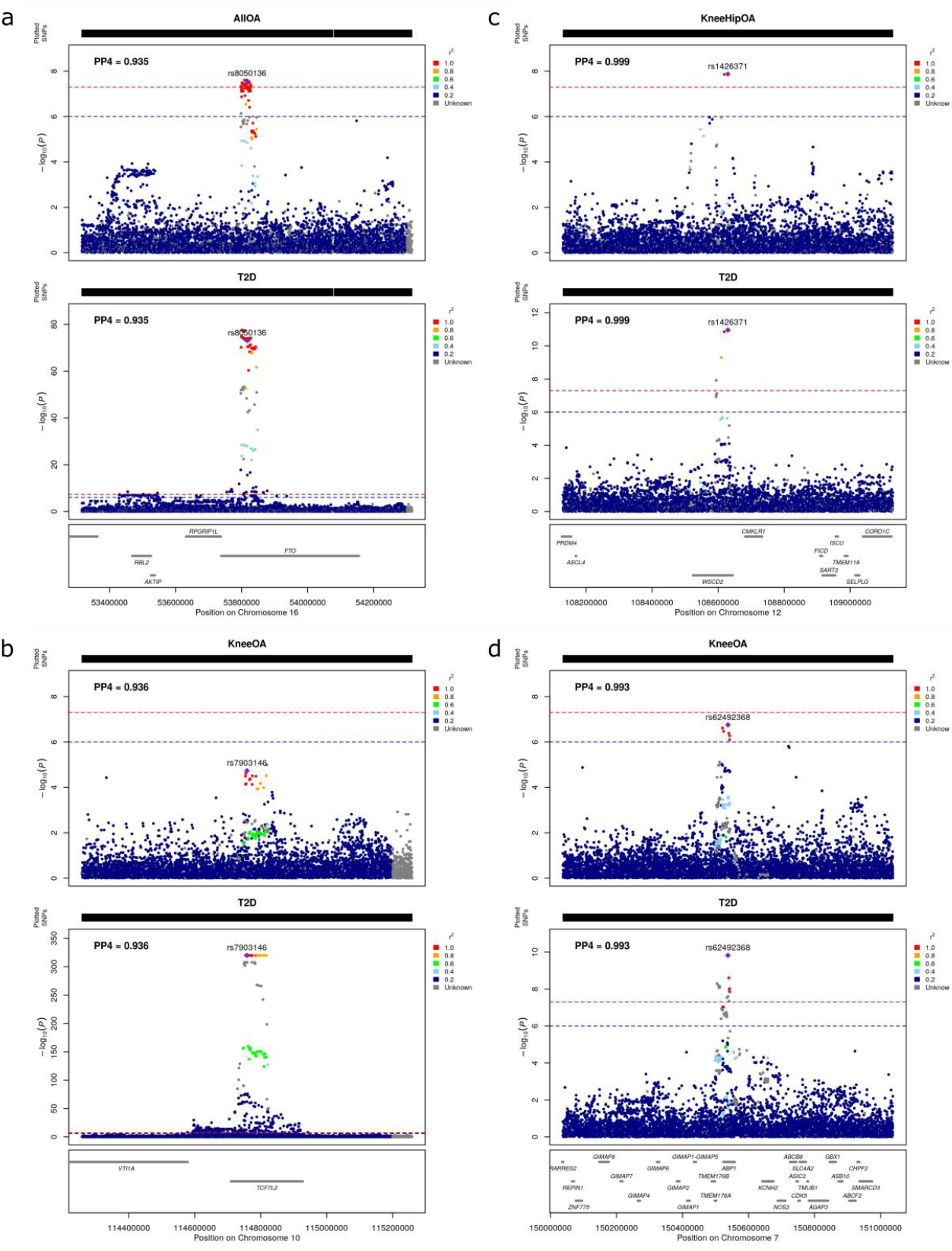
Regional association plots of the highlighted colocalizing regions between type 2 diabetes and osteoarthritis. a) FTO and IRX3 region; b) TCF7L2 region; c) WSCD2 and TMEM119 region; d) TMEM176A region. The plots are colored based on linkage disequilibrium between the lead causal variant and all other variants. (PP4 = posterior probability of a single shared causal variant; OA = osteoarthritis; T2D = type 2 diabetes; All OA = osteoarthritis at any site)

*FTO* is a high-confidence osteoarthritis effector gene involved in skeletal development, adipogenesis, and neuronal function and development [23]. It is also associated with syndromic human disease *growth retardation, developmental delay, and facial dysmorphism (GDFD)*, which is a lethal autosomal recessive multiple congenital anomaly syndrome [24]. *IRX3* is a known functional long-range target of *FTO* variants associated with obesity [25]. *FTO* and *IRX3* knockout mice show decreased body weight, decreased bone mineral density, and improved glucose tolerance (high bone mineral density is a risk factor for hip and knee osteoarthritis [26]). We expected genes with an adiposity-driven effect to be involved in the shared genetic aetiology of the type 2 diabetes-osteoarthritis comorbidity since obesity constitutes a common risk factor [8].

#### TCF7L2

*TCF7L2* is one of the highest-scoring effector genes. This genomic locus colocalizes for type 2 diabetes and knee osteoarthritis with a posterior probability of a shared causal variant of over 93% (Figure 5). Here, the 95% credible set for the causal variant from the colocalization analysis consists of three variants, which have opposite risk-increasing alleles for type 2 diabetes and knee osteoarthritis (Table S8A). *TCF7L2* has not been associated with osteoarthritis at genome-wide significance levels to date (all variants in the credible set achieve nominal significance in the latest knee osteoarthritis GWAS [23]). Genetic variants associated with *TCF7L2* expression in pancreatic islets and in osteoarthritis cartilage colocalize with this association signal. For the variants in both 95% credible sets from these colocalization analyses, the risk-increasing alleles for type 2 diabetes are associated with a lower BMI, a lower risk of knee osteoarthritis and an increased expression of *TCF7L2* in pancreatic islets and osteoarthritic cartilage.

We find evidence that increased BMI causes decreased expression of *TCF7L2* in intact and degraded cartilage (Table S7A). Additionally, we find evidence that an increase in *TCF7L2* expression in pancreatic islets causes an increase in type 2 diabetes risk (OR=5.1; 95%CI=[4.7, 5.4]; p-value<1E-300; F-stat=2018) and a decrease in knee osteoarthritis risk (OR=0.83; 95%CI=[0.77, 0.91]; p-value=1.77E-05; F-stat=18.4). These results are in line with the evidence shown above in support of an opposite effect of the genetic variants associated with the expression of *TCF7L2* in knee osteoarthritis and type 2 diabetes risk.

*TCF7L2* is among the leading signals for type 2 diabetes risk, persisting following adjustment for BMI [27]. Our results suggest that *TCF7L2* exerts an effect that goes through an alternative biological pathway to increased BMI. It has been shown that isoforms of *TCF7L2* regulate the expression of genes related to cartilage destruction in human chondrocytes [28]. *TCF7L2* is a key effector gene of the Wnt/β-catenin signalling pathway. This pathway plays a role in both type 2 diabetes, through glucose homeostasis, and in osteoarthritis, through cartilage and bone formation [29], [30].

#### TMEM119 and WSCD2

Two high-confidence effector genes, *WSCD2* and *TMEM119*, reside in the same genomic locus, which colocalizes for type 2 diabetes and knee osteoarthritis with a posterior probability of 99.9% (Figure 5). The 95% credible set consists of two variants: rs1426371 and rs3764002, an intronic and a missense variant (amino acid change: Thr266Ile) within *WSCD2*, respectively. The risk-increasing alleles of both variants are concordant for osteoarthritis of the knee and type 2 diabetes. The variant with the highest posterior probability of being causal for osteoarthritis and type 2 diabetes, rs1426371, has reached genome-wide significance levels in the latest knee osteoarthritis GWAS meta-analysis [23]. The missense variant, rs3764002, is associated with BMI, type 2 diabetes, lean mass, and neuropsychiatric disorders including anxiety and schizophrenia. The missense variant is predicted to alter protein function and this alteration is predicted to be damaging by both SIFT (https://sift.bii.a-star.edu.sg) and PolyPhen (http://genetics.bwh.harvard.edu/pph2/).

*WSCD2* eQTLs in degraded osteoarthritis cartilage colocalize with type 2 diabetes and knee osteoarthritis with a posterior probability of 99%. The lead eQTLs are rs142637 and rs3764002. The expression level-increasing alleles are the same as the risk-increasing alleles for both diseases (Table S8B). *WSCD2* is also a differentially expressed gene (DEG) in pancreatic islets from diabetes patients versus healthy controls and is downregulated in diabetic islets. Moreover, it has been previously shown that *WSCD2* is functionally associated with type 2 diabetes and positively correlated to insulin secretion [31],[32]. Conclusions from causal inference analysis were limited due to weak instruments (F-statistic<10), which can bias causal effect inference (Table S7). Further research is needed to better understand the biological mechanisms through which *WSCD2* influences the type 2 diabetes-osteoarthritis comorbidity.

*TMEM119* is the second most highly scoring high-confidence effector gene. It is a DEG in osteoarthritis cartilage and pancreatic islets and is more highly expressed in degraded compared to intact cartilage and in healthy compared to pancreatic islets. Knockout mice for *TMEM119* show phenotypes related to both osteoarthritis and type 2 diabetes, such as decreased body weight, impaired osteoblast differentiation, and decreased compact bone thickness. *TMEM119* is related to bone formation by promoting osteoblast differentiation [33]. Fewer osteoblasts can lead to a decrease in compact bone thickness, which is also observed in knockout mice [34]. The over-expression of *TMEM119* in degraded cartilage from osteoarthritis patients supports the evidence of an increase in bone formation in later stages of the disease [35]. However, the lower expression of *TMEM119* in diabetes compared to healthy pancreatic islets points to further potential mechanisms of effect in the comorbidity.

#### TMEM176A

*TMEM176A*, also a high-confidence effector gene, has not been previously identified as implicated in either osteoarthritis or type 2 diabetes. This locus colocalizes for type 2 diabetes and knee osteoarthritis with a posterior probability of a shared causal variant of 99.3% (Figure 5). The index variants are rs62492368 and rs7794796, both located in the intron of the *AOC1* gene. rs62492368 is associated with type 2 diabetes [36] and rs7794796 is associated with appendicular lean mass [37]. Type 2 diabetes and osteoarthritis show opposite risk-increasing alleles for all variants in the 95% credible set from the colocalization analysis (Table S8C). These variants colocalize with PP4 > 0.8 between the diseases and eQTL data from pancreatic islets and synovium. The index variants from the colocalization with eQTLs also have opposite risk-increasing alleles for both diseases. Similarly, to the *TCF7L2* locus case, our results suggest that the mechanism through which *TMEM176A* exerts an effect on osteoarthritis and type 2 diabetes may have contrary directions.

We found body fat percentage to be linked to a decrease in the expression of *TMEM176A* in synovium (Table S7A), albeit with weak instruments (F-statistic<10). Decreased expression of *TMEM176A* in intact osteoarthritis cartilage and pancreatic islets was associated with reduced risk of type 2 diabetes (pancreatic islets: OR=1.14, 95%CI=[1.07, 1.21], p-value=8.5E-05, F-stat=15.4; cartilage: OR=1.05, 95%CI=[1.03, 1.08], p-value=8.5E-05, F-stat=15.4) and increased risk of knee osteoarthritis (pancreatic islets: OR=0.93; 95%CI=[0.87, 0.99]; p-value=0.048; F-stat=3.9, cartilage: OR=0.97; 95%CI=[0.94, 0.99]; p-value=0.048; F-stat=3.9). This genomic locus is one of the two colocalizing regions that do not show any evidence of statistical colocalization between type 2 diabetes, osteoarthritis and the adiposity measures studied here. This suggests that this region, and possibly *TMEM176A*, acts through an alternative biological path to adiposity.

Results of the causal inference analysis mirror the output of differential expression analyses conducted in pancreatic islets and osteoarthritic cartilage [38], [39]. While *TMEM176A* is more highly expressed in diabetes compared to healthy pancreatic islets, an increase of its expression in the same tissue has a causal effect on increased risk of type 2 diabetes. Similarly, while *TMEM176A* was found to have lower expression in degraded compared to intact cartilage, the reduced expression of this gene in intact cartilage has a causal effect on increased risk of total knee replacement.

## Discussion

We present an approach to disentangle the shared genetic aetiology between two co-occurring chronic diseases, exemplified through the type 2 diabetes-osteoarthritis comorbidity. Studies have shown a stronger association of BMI with osteoarthritis of the knee than of the hip [12]. We find stronger statistical evidence of a genetic correlation between type 2 diabetes and knee osteoarthritis compared to hip osteoarthritis. By leveraging the latest large-scale GWAS for both diseases, we find robust evidence of colocalization at 18 genomic loci and, by incorporating multi-omics and functional genomics information, we derive a list of 19 high-confidence effector genes for the comorbidity. The majority of genomic loci colocalize for type 2 diabetes and knee, rather than hip, osteoarthritis, in keeping with the genome-wide correlation analysis results.

Our findings support the epidemiological link between obesity, osteoarthritis, and type 2 diabetes. In this case, only two of the 18 colocalized regions do not colocalize with measures of adiposity. Several of the high-confidence genes are associated with obesity-related traits, including *FTO* and *IRX3*. We show that the identified high-confidence effector genes are enriched for biological pathways associated with adiposity. Stratifying the high-confidence effector genes into knee or hip osteoarthritis provides further insight into the biological mechanisms underlying the comorbidity. High-confidence effector genes mostly related to hip osteoarthritis are also enriched for biological pathways of skeletal formation, which underlines the strong link between bone development and hip osteoarthritis [7]. Given that two-thirds of FDA-approved drugs are supported by genetic evidence, we explore the druggable potential of the prioritized genes [40]. We highlight approved drugs currently used for the treatment of diabetes, obesity, pain, and inflammation.

Observational studies report that the positive association between type 2 diabetes and osteoarthritis persists after adjusting for BMI [9]. As BMI only captures a limited subset of the effect of adiposity on the comorbidity, this could be a source of residual confounding due to measurement error [41] and observed attenuation of association can be underestimated. We performed in-depth analyses to disentangle the role of adiposity on the comorbidity and find evidence that *TCF7L2* and *TMEM176A* exert an effect on type 2 diabetes and osteoarthritis through an alternative biological path. Further examination, including functional studies, is needed to dissect the precise way in which these genes affect the comorbidity.

Type 2 diabetes and insulin resistance are known to be negatively correlated with bone strength and are also associated with bone fracture [42]. One possible link between bone and lipid metabolism is the fact that osteoblasts and adipocytes share a common progenitor cell in adult bone marrow with a degree of plasticity that can lead to an imbalance between the two cell lineages [43]. In support of this link, differentiation regulation of osteoblasts is highlighted by one of the identified high-confidence effector genes, *TMEM119*. In summary, we highlight three potential biological mechanisms underpinning the comorbidity between type 2 diabetes and osteoarthritis: obesity, imbalance between osteoblasts and adipocytes differentiation in adult bone marrow and the Wnt/β- catenin signalling pathway.

The genetic and functional genomic data employed in this study are biased towards European populations. Going forward, it will be important to expand analyses to data from diverse populations. The eQTL data from pancreatic islets used in the analyses here comprise almost four times as many samples as the eQTL data from chondrocytes. Therefore, molecular QTL analyses for type 2 diabetes-relevant tissues were better-powered than for osteoarthritis-relevant tissues. Mendelian randomization and the subsequent sensitivity analyses were conducted within the constraints of available instruments for expression of the high-confidence genes. This was partially because the molecular QTL data used in this work include cis-QTLs only, which restricts the analyses to variants within the vicinity of the genes or proteins of interest. Future studies should include a wider array of (as yet unavailable) genome-wide molecular QTLs, including at the single-cell level.

We have studied one of the most frequently co-occurring pairs of complex diseases: type 2 diabetes and osteoarthritis. Our findings offer insights into the biological processes underpinning the comorbidity and highlight potential drug repurposing opportunities in addition to new targets. As the world population life expectancy continues on an upward trajectory, the challenge of tackling multimorbidity will continue to be high on the healthcare agenda. Genomic data-based approaches, as exemplified here for type 2 diabetes and osteoarthritis, can help improve our understanding of the co-occurrence of chronic conditions.

## Data Availability

- This paper analyzes existing, publicly available data. These accession numbers for the datasets are listed above.
- All original code has been deposited at Zenodo and is publicly available as of the date of publication. https://doi.org/10.5281/zenodo.7525171
- Any additional information required to reanalyze the data reported in this paper is available from the lead contact upon request.

https://msk.hugeamp.org/downloads.html

https://www.diagram-consortium.org/

https://portals.broadinstitute.org/collaboration/giant/index.php/GIANT_consortium

http://www.nealelab.is/uk-biobank/

https://msk.hugeamp.org/downloads.html

https://zenodo.org/record/3408356

https://t2d.hugeamp.org/

## Acknowledgements

We thank Lorraine Southam and Konstantinos Hatzikotoulas for their input and support. We also thank the Genetics of Osteoarthritis (GO), the DIAMANTE and the InsPIRE consortia for providing their data.

## Author contributions

E.Z. and A.L.S.V.A conceived and designed the study and wrote the manuscript. A.L.S.V.A and A.H. performed computation and statistical analyses and produced the figures. A.L.S.V.A, G.D.S, A.H., E.Z. and A.P.M. performed data interpretation. A.L.S.V.A, G.D.S, A.H., E.Z. G.K, and A.P.M. provided significant feedback on the methods, results, and manuscript.

## Data and code availability

- This paper analyzes existing, publicly available data. These accession numbers for the datasets are listed in the key resources table.
- All original code has been deposited at Zenodo and is publicly available as of the date of publication. DOIs are listed in the key resources table.
- Any additional information required to reanalyze the data reported in this paper is available from the lead contact upon request.

## Methods

### Resource availability

#### Lead contact

Further information requests should be directed to the lead contact, Eleftheria Zeggini.

#### Materials availability

This study did not generate new unique reagents.

### Methods details

#### Datasets

For osteoarthritis, we used the largest GWAS meta-analysis to date, from the Genetics of Osteoarthritis (GO) consortium [23]. In total, it comprises data from 826,690 individuals (177,517 cases) from mostly white European ancestry for 11 different osteoarthritis phenotypes. In this study, we used following osteoarthritis phenotypes: knee, hip, knee and/or hip, total knee replacement (TKR), total hip replacement (THR), total joint replacement (TJR), and osteoarthritis at any site (all). A table with the number of cases, controls, and the total patients for each study can be found in Table 1. For type 2 diabetes, the GWAS meta-analysis unadjusted for BMI from the DIAMANTE consortium was used [36]. It includes data from 898,130 individuals (74,124 cases) of European ancestry.

**Table 1:** an overview of osteoarthritis phenotypes used in this work and the number of cases, controls and the total number of patients included in the corresponding GWAS.

| OA phenotype | Cases | Controls | Total |
| --- | --- | --- | --- |
| All | 177,517 | 649,173 | 826,690 |
| Knee | 62,497 | 333,557 | 396,054 |
| Knee and/or hip | 89,741 | 400,604 | 490,345 |
| Hip | 36,445 | 316,943 | 353,388 |
| TKR | 18,200 | 233,841 | 252,041 |
| TJR | 40,887 | 327,689 | 368,576 |
| THR | 23,021 | 296,016 | 319,037 |

We also employed molecular QTL data from disease-specific tissues. For osteoarthritis, we used eQTL data from intact cartilage (n=95), degenerated cartilage (n=87) and synovium (n=77), as well as pQTL data from intact and degenerated cartilage (n=99) [44]. All samples were collected from osteoarthritis patients. For type 2 diabetes, we used eQTL data from pancreatic islets from the InsPIRE consortium [45]. In the pancreatic islets data set, 37 patients out of 420 were diabetic.

We aligned the effect alleles of all data sets used in this paper by inverting the sign of the effect sizes when a mismatch was detected. Chromosome X was not included in any analysis. All data sets used the Genome Reference Consortium Human Build 37 (GRCh37) assembly.

##### Measures of adiposity

We used four measures of adiposity: body mass index (BMI), waist-to-hip ratio (WHR) unadjusted for BMI, whole body fat mass and body fat percentage. For BMI (N=806,834) and WHR (N=697,734), we used the latest meta-analysis combining data from the GIANT consortium and the UK biobank [54]. The inverse rank normalized GWAS summary statistics for whole body fat mass (N=330,762) and body fat percentage (N=331,117) were taken from the Neale’s Lab website (http://www.nealelab.is/uk-biobank/). For each adiposity phenotype, we looked up the effect and significance of all variants in the 95% credible set of the colocalized regions between type 2 diabetes and osteoarthritis (Table S9).

### Quantification and statistical analysis

#### Genetic overlap of type 2 diabetes and osteoarthritis phenotypes

We conducted a linkage disequilibrium (LD) score regression analysis using the LDSC software (v1.0.1) with –rg flag to estimate the genetic correlation between each osteoarthritis phenotype and type 2 diabetes (Table S1) [46]. Since the majority of the GWAS used here comprises data of European ancestry individuals only, pre-computed LD scores from the 1000 Genomes European ancestry haplotypes were used [47]. To assess the potential for chance findings when performing multiple statistical analyses, we performed a permutation-based analysis. We randomly permuted the effects (Z-scores) of the variants for the osteoarthritis phenotypes ten thousand times while fixing the effects for type 2 diabetes. Running LD score regression on each permuted data set yielded an empirical p-value for the genetic correlation of type 2 diabetes and each analysed osteoarthritis phenotype.

#### Statistical colocalization analysis

We defined regions of 2 Mb (+-1 Mb) around established independent association signals from each disease. For type 2 diabetes, we selected all primary and secondary independent signals from the BMI unadjusted GWAS (p-value threshold = 5*x*10^−8^). For osteoarthritis, we selected the risk signals for the respective phenotype at the adjusted genome-wide significance of 1.3*x*10^−8^. For each osteoarthritis phenotype, we performed regional pairwise statistical colocalization analysis with type 2 diabetes using the *coloc*.*abf* function from the *coloc* R package (version 3.2.1) [48]. Colocalization analyses were conducted using estimated regression coefficients (effect sizes) and standard errors (Table S2). In short, this function calculates posterior probabilities for five association configurations under the assumption of a single causal variant per trait. These configurations are summarized in the hypotheses below:

***H0:*** no trait has a genetic association in the region

***H1:*** trait 1 has a genetic association in the region

***H2:*** trait 2 has a genetic association in the region

***H3:*** both traits have a genetic association in the region, but with different causal variants

***H4:*** both traits share a genetic association (single causal variant) in the region

For all osteoarthritis phenotypes, we used the default prior probabilities of the *coloc* R package. We considered evidence for colocalization if the posterior probability of H4 (PP4) > 0.8. For each genomic locus of colocalization, we calculated a 95% credible set for the causal variant by taking the cumulative sum of the variants’ posterior probabilities to be causal conditional on H4 being true. LD between the single nucleotide polymorphisms (SNPs) was calculated using plink (version 2.0 alpha) [49] based on the UK biobank [50] and was used for visualizing the results in regional association plots.

#### Knockout mouse phenotypes

We performed a schematic search for each gene in the vicinity of the colocalized genomic loci that colocalize between type 2 diabetes and osteoarthritis to screen for knockout mice showing phenotypes related to type 2 diabetes or osteoarthritis. The databases used in this scope were the International Mouse Phenotyping Consortium (IMPC) (https://www.mousephenotype.org/), Mouse Genome Informatics (MGI) (http://www.informatics.jax.org/) and Rat Genome Database (RGD) (https://rgd.mcw.edu/) databases. For IMPC and RGD we extracted the knockout mice phenotypes for each potential effector gene using the programmatic data access via their application programming interface (API). For MGI, we used the MGI batch query.

For type 2 diabetes, we looked for insulin and diabetes-related phenotypes that included the following terms: insulin, glucose, diabetes, hyperglycaemia, pancreas, pancreatic, obesity, BMI, body weight, body mass, body fat, beta cell, and glucosuria. For osteoarthritis, we looked for musculoskeletal phenotypes including the terms skeletal, muscle, bone, osteo, arthritis, muscular, joint, body size, growth, stature, and height.

#### Rare and syndromic human diseases

To investigate whether any analysed genes are associated with a monogenic disorder, we extracted data from the Online Mendelian Inheritance in Man (OMIM) (https://omim.org/) using their API. The terms we looked up for osteoarthritis-related phenotypes were bone, muscle, skeleton, osteo, arthritis, muscular, joint, body size, growth, skeletal, stature, height, Hand-foot-uterus, synostosis, Martsolf, Warburg, leukodystrophy, squalene, and Finca. For type 2 diabetes we searched for insulin, glycemia, glucose, diabetes, pancreas, pancreatic, obesity, BMI, body weight, body mass, body fat, beta cell, glucosuria, Martsolf, aciduria, Aicardi-Goutières, and Finca.

#### Differential gene expression

We explored if the analysed genes show differential expression for type 2 diabetes and osteoarthritis using published summary statistics from RNA-seq datasets. For osteoarthritis, differential expression was assessed by comparing paired intact and degraded osteoarthritis cartilage from 124 patients [38]. Since the samples were collected within patients, the data is automatically robust against cofactors such as age and population structure. For type 2 diabetes, we used RNA-seq data from surgical pancreatic tissue samples from metabolically phenotyped pancreatectomized patients. Samples were collected from 18 non-diabetic patients and 39 patients that were previously diagnosed with type 2 diabetes [39]. The differential expression analysis was based on a linear model with age, sex, and BMI as covariates. We considered genes that changed more than 1.5-fold in either direction and had an adjusted p-value < 0.05 to be differentially expressed between degraded (high-grade) and intact (low-grade) osteoarthritis cartilage, and diabetic versus healthy pancreatic islets, for osteoarthritis and type 2 diabetes respectively.

#### Multi-trait statistical colocalization analysis with eQTL and pQTL data

First, we superimposed molecular QTL information from disease-specific tissues by performing multi-trait molecular QTL-GWAS colocalization analyses. The analyses were performed only on the variants in the 95% credible set. The input consisted of three summary statistics: one from the type 2 diabetes GWAS, one from the osteoarthritis phenotype GWAS, and one from the disease-relevant tissue molecular QTL data set. Since one variant is tested for multiple genes in an eQTL data set, or multiple proteins for the pQTL data sets, we performed the colocalization gene-wise or protein-wise, respectively, such that for each analysis a single molecular QTL summary statistic is available for each variant. If the 95% credible set consisted only of a single variant, for each gene or protein, we included all variants in a 1 Mb window in the analysis.

For the multi-trait statistical colocalization analyses, we used the R package *HyPrColoc* (version 1.0.0) [51]. We conducted regional gene-wise analysis to assess whether all traits colocalize by switching off the Bayesian divisive clustering algorithm (bb.alg=FALSE). In a similar manner to the *coloc* package, we used *HyPrColoc* to estimate the posterior probabilities and identify candidate causal genes using multiple traits as input. For consistency, evidence for colocalization was considered at a threshold of 0.8 for PP4. The type 2 diabetes and osteoarthritis GWAS meta-analyses share five cohorts. Although the sample overlap, we assumed independence between the data sets, as instructed by the developers of the *HyPrColoc* package.

Using the prior knowledge that type 2 diabetes and osteoarthritis colocalize in the analysed genomic loci, we adapted the prior parameters of the *HyPrColoc* algorithm accordingly. The first parameter, *prior*.*1*, which denotes the probability of a SNP being associated with one trait only, was set to 1e-10, six times smaller than the default. We set the second parameter *prior*.*2* to 0.7 instead of the default of 0.98. 1-*prior*.*2* denotes the prior probability of a SNP being associated with an additional trait and 1-(*prior*.*2*)^2^ with the SNP being associated with the two other traits. LD between SNPs was again calculated using plink (version 2.0 alpha) [49] based on the UK biobank [50].

#### Scoring of potential effector genes

In genomic loci that colocalized between type 2 diabetes and at least one osteoarthritis phenotype with a PP4 > 0.8, we analysed all genes in a 1 Mb window on either side of the lead variant of the 95% credible set. We incorporated orthogonal multi-omics and functional information to derive a list of high-confidence effector genes for the type 2 diabetes-osteoarthritis comorbidity.

Except for the pQTL analysis, all four above-mentioned biological lines of evidence were tested for both osteoarthritis and type 2 diabetes, yielding one separate score for each disease. Additionally, we incorporated information about previously established high-confidence effector genes for the individual diseases. For type 2 diabetes, we defined genes as high-confidence if their top score in the type 2 diabetes knowledge portal was at least 4 (https://t2d.hugeamp.org/). For osteoarthritis we selected genes scored as high-confidence by the GO consortium [23]. Since our analysis overlaps with criteria used to define a gene as high-confidence for the individual diseases, we followed an approach to incorporate this information orthogonally: if a gene is high-confidence for a disease, but scored zero in our analysis, we updated the respective disease score to one.

We also looked up all variants in the 95% credible sets searching for any missense variants. This look-up was summarized in an additional score, the missense variant score. The total score was defined as the sum of the osteoarthritis score, the type 2 diabetes score, and the missense variant lookup. If, however, for a gene only the missense variant score is non-zero, the total score was set to zero since it is not relevant for the type 2 diabetes-osteoarthritis comorbidity.

Based on the scoring of the six orthogonal biological lines of evidence, we defined genes as potential effector genes if they showed at least one line of evidence for either one of the diseases. Genes that scored at least one line of evidence for osteoarthritis and one for type 2 diabetes were defined as likely effector genes for the comorbidity. High-confidence effector genes were a subset of the likely effector genes that scored at least 3 in the total score (Table S3).

To further analyse our set of effector genes, we grouped them according to the osteoarthritis localisation. If a gene is only associated with osteoarthritis at any site, knee and/or hip osteoarthritis and TJR, then it was considered to be associated with both knee and hip osteoarthritis. If, in addition, an association with knee and/or TKR is observed, then we consider the gene to be mostly associated with knee osteoarthritis. Similarly, if it is associated with hip or THR, then the gene is classified as mostly related to hip osteoarthritis.

#### Multi-trait statistical colocalization analysis with adiposity measures

In the genomic regions that colocalized between type 2 diabetes and osteoarthritis, we performed multi-trait colocalization analyses between type 2 diabetes, osteoarthritis and the above-mentioned measures of adiposity. The analyses were performed only on the variants in the 95% credible set. If the 95% credible set consisted only of a single variant, for each gene or protein, we included all variants in a 1 Mb window on either side of the single variant in the analysis. As for the molecular QTL colocalization, we used the same functions of the R package *HyPrColoc* (version 1.0.0) and adjusted the prior parameters accordingly (*prior*.*1*=1e-10, *prior*.*2*=0.7) [51]. For consistency, evidence for colocalization was considered at a threshold of 0.8 for PP4.

#### Pathway analysis

We performed gene set enrichment analyses on the likely and on the high-confidence effector genes stratified by knee or hip osteoarthritis association (Table S5). The number of genes in each set is summarized in Table 2. We used the human resources and the enrichment software from the ConsensusPathDB (http://cpdb.molgen.mpg.de/) to examine the functional annotation of each gene set by testing their enrichment among curated networks in humans [52]. We used the networks from Reactome, KEGG, WikiPathways and Gene Ontology. For the latter, we included the subcategories molecular function, biological processes, and cellular component up to level 4. We required a minimum overlap of 2 genes for enrichment. The significance threshold was set at FDR < 0.05.

**Table 2:** number of genes in each gene set (HC =high-confidence)

| Gene set | Number of genes |
| --- | --- |
| Likely effector genes | 72 |
| Likely effector genes related to knee | 67 |
| Likely effector genes related to hip | 43 |
| HC effector genes | 19 |
| HC effector genes related to knee | 18 |
| HC effector genes related to hip | 10 |

#### Druggable genome

To outline drug repurposing targets, we queried the druggability status of the 72 likely effector genes for the comorbidity. We used the Druggable Genome database, which consists of 4479 genes that are classified into three tiers depending on their progress in the drug development pipeline [15]. Tier 1 included 1427 genes that are clinical-phase drug candidates or targets of already approved small molecules and biotherapeutic drugs. Tier 2 consisted of 682 genes that encode targets with known bioactive drug-like small-molecule binding partners and genes with ≥ 50% identity (over ≥75% of the sequence) with approved drug targets. Tier 3 comprised 2370 genes encoding secreted or extracellular proteins, proteins with more distant similarity to approved drug targets, and members of key druggable gene families that were not included in tier 1 or 2. Tier 3 was further subdivided to prioritize genes in proximity (+-50 kbp) to a GWAs SNP from the GWAS catalog and had an extracellular location (Tier 3A). Tier 3B is composed of the remaining genes.

For the likely effector genes included in tier 1, we further examined the approved or in clinical trial drugs using the DrugBank online database (https://www.drugbank.com, accessed on the 1^st^ of August 2022).

#### Causal inference analysis

Causal inference was strengthened through use of bi-directional two-sample Mendelian randomization (MR) between type 2 diabetes and all analysed osteoarthritis phenotypes [10]. We used the *TwoSampleMR* R package (version 0.5.6), which is curated by MR-Base [53]. We performed causal inference analyses on the full summary statistics (Table S6). For all analyses, instrumental variables (IVs) were selected as the genome-wide significant (p-value ≤ 5*x*10^−8^) and independent SNPs from the full data. Independence was defined as LD-based clumped SNPs with a strict LD threshold of *R*^2^ = 0.001 over a 10Mb window on either side of the index variant. To assure that the IVs are more strongly related to the exposure than to the outcome, we applied Steiger filtering [10]. We applied the inverse variance weighted (IVW) method, which performs a random-effects meta-analysis of the Wald ratios for each SNP and the weighted median (WM) method. Finally, we performed sensitivity analyses by testing for heterogeneity based on the Q-statistic using the *mr_heterogeneity* function from the *TwoSampleMR* R package. Horizontal pleiotropy was assessed through the intercept of the MR-Egger regression. To account for multiple testing, p-values were adjusted using the false discovery rate (FDR) approach [10].

#### Two-step Mendelian randomization

We performed a two-step Mendelian randomization analysis between different adiposity measures and type 2 diabetes or osteoarthritis using *cis* expression QTLs (eQTLs) of each high-confidence effector gene in disease-relevant tissues as mediators (Table S7) [55]. In the first step, we assess whether adiposity is causal for the expression of our genes in the respective analysed tissues. To assure independence of IVs between the two steps, we excluded independent eQTLs from the risk variants of each adiposity measure. Independence was defined by local LD-based clumping with *R*^2^ = 0.001 over a 10Mb window on either side of the index variant.

For the second step, we used independent genetics variants associated to each high-confidence gene as IVs and conducted a two-sample MR analysis between each of our genes and type 2 diabetes or osteoarthritis. For each analysed tissue and each gene-disease pair, we conducted one MR analysis using the *TwoSampleMR* R package (version 0.5.6) [53]. If only one SNP was available after clumping and harmonizing the data, we employed the Wald ratio method. If more than one SNP remained after the pre-processing steps, we applied the IVW method and tested for heterogeneity with the *mr_heterogeneity* function. Moreover, if more than three SNPs were used for the causal inference analysis, we also tested for horizontal pleiotropy through MR-Egger regression. Additionally, we estimated the F-statistics from summary level data as *mean(beta*^2^*/se*^2^) to assess the strength of the IVs [10]. Finally, we adjusted the p-values for multiple testing using the FDR approach.

#### Tissue fractionation

We determined the tissue-specific role of BMI in both osteoarthritis and type 2 diabetes using MR restricted to BMI instruments colocalizing with eQTLs in brain and adipose tissue, respectively, as described in [22] (Table S10). Briefly, summary-level MR was performed restricted to the 86 adipose tissue colocalizing SNPs and the 140 brain tissue colocalizing SNPs, where the numerator of the Wald ratio is the SNP effect on osteoarthritis or type 2 diabetes and the denominator is the effect estimate for the SNP on BMI from a GWAS meta-analysis of UK Biobank and the GIANT consortium [56] available at the MR-Base platform [53]. Osteoarthritis summary statistics were extracted from the latest GWAS of hip and knee osteoarthritis from the GO consortium [23]. Type 2 diabetes summary statistics were extracted from the latest European DIAMANTE consortium GWAS [36].

We used inverse variance weighted meta-analysis of the individual SNP Wald ratios to estimate the causal effects of adipose tissue-instrumented BMI and brain tissue-instrumented BMI on each outcome. As sensitivity analyses, we performed MR-Egger to determine the potential role of pleiotropic effects (i.e. mediated via BMI-independent pathways), which gives a pleiotropy robust estimate of the causal effect assuming that there is no correlation between instrument strength (i.e. the association of the SNP with BMI) and the pleiotropic effect [10]. We also performed weighted median analysis, which gives an unbiased estimate of the causal effect as long as less than 50% of the SNPs are invalid instruments [57]. We performed a z-test to assess the effect difference between adipose and brain tissue-instrumented BMI MR analyses.

## Supplemental information titles

**Table S1**: Genetic correlation matrix for type 2 diabetes and osteoarthritis.

**Table S2**: Overview of colocalized genomic loci between type 2 diabetes and osteoarthritis with additional information about colocalization with molecular QTLs from disease-relevant tissues.

**Table S3A**: Scoring of all genes in a 1Mb window on either side of the lead variant of the 18 colocalized regions between type 2 diabetes and osteoarthritis.

**Table S3B**: Overview of scoring of likely effector genes for the type 2 diabetes and osteoarthritis comorbidity.

**Table S3C**: Overview of scoring of high confidence effector genes for the type 2 diabetes and osteoarthritis comorbidity.

**Table S4**: Likely effector genes for the type 2 diabetes and osteoarthritis comorbidity included in the druggable genome.

**Table S5A**: Results of the pathway analyses of the high confidence effector genes for the type 2 diabetes-osteoarthritis comorbidity stratified by knee or hip association.

**Table S5B**: Results of the pathway analyses of the likely effector genes for the type 2 diabetes-osteoarthritis comorbidity stratified by knee or hip association.

**Table S6A**: Results of bidirectional two-sample Mendelian randomization analyses between type 2 diabetes and osteoarthritis.

**Table S6B**: Steiger-filtered results of bidirectional two-sample Mendelian randomization analyses between type 2 diabetes and osteoarthritis.

**Table S7A**: First step results of two-step Mendelian randomization analysis between adiposity measures and gene expression of high confidence effector genes for the type 2 diabetes and osteoarthritis comorbidity in disease-relevant tissues.

**Table S7B**: Second step results of two-step Mendelian randomization analysis between gene expression of high confidence effector genes for the type 2 diabetes and osteoarthritis comorbidity in disease-relevant tissues and either one of the diseases.

**Table S9A**: Multi-trait colocalization between type 2 diabetes, osteoarthritis and BMI for regions that colocalized between type 2 diabetes and osteoarthritis.

**Table S9B**: Direction of effect and p-value of variants in the 95% credible set of the colocalized regions for the comorbidity.

**Table S10**: Results of two-sample Mendelian randomization analyses between type 2 diabetes or osteoarthritis and BMI risk variants associated with eQTLs in brain and subcutaneous adipose tissue.

