## Supplemental Figures for "Insights into the comorbidity between type 2 diabetes and osteoarthritis"

| | rs7903146 | | | rs35198068 | | | rs34872471 | | |
|---|---|---|---|---|---|---|---|---|---|
| | EA | beta | pval | EA | beta | pval | EA | beta | pval |
| Knee OA | T | -0.035 | 1.8e-5 | T | 0.035 | 2.4e-5 | T | 0.034 | 3.2e-5 |
| T2D | T | 0.31 | < 1e-300 | T | -0.31 | < 1e-300 | T | -0.31 | < 1e-300 |

**Table S8A**: Overview of effect size of the variants in the 95% credible set of the *TCF7L2* region for type 2 diabetes and osteoarthritis.

| | rs1426371 | | | rs3764002 | | |
|---|---|---|---|---|---|---|
| | EA | beta | pval | EA | beta | pval |
| **Knee OA** | A | -0.0515 | 8.86E-10 | T | -0.0508 | 8.90E-10 |
| **Knee and/or hip OA** | A | -0.0408 | 1.34E-08 | T | -0.0402 | 1.39E-08 |
| **All OA** | A | -0.0246 | 3.62E-06 | T | -0.0242 | 3.77E-06 |
| **TJR** | A | -0.0412 | 2.67E-05 | T | -0.0403 | 3.27E-05 |
| **T2D** | A | -0.05 | 1.10E-11 | T | -0.049 | 1.40E-11 |

**Table S8B:** Overview of effect size of the variants in the 95% credible set of the *WSCD2* and *TMEM119* region for type 2 diabetes and osteoarthritis.

| | rs62492368 | | | rs7794796 | | |
|---|---|---|---|---|---|---|
| | **EA** | **beta** | **pval** | **EA** | **beta** | **pval** |
| **Knee OA** | A | -0.0397 | 1.77E-07 | T | -0.1132 | 0.01828 |
| **Knee and/or hip OA** | A | -0.0303 | 3.96E-06 | T | -0.0281 | 1.26E-05 |
| **TKR** | A | -0.074 | 2.17E-08 | T | -0.0769 | 2.88E-09 |
| **TJR** | A | -0.04 | 1.58E-05 | T | -0.0404 | 8.06E-06 |
| **T2D** | A | 0.044 | 1.50E-10 | T | 0.04 | 2.50E-09 |

**Table S8C**: Overview of effect size of the variants in the 95% credible set of the *TMEM176A* region for type 2 diabetes and osteoarthritis.

*T2D = type 2 diabetes; OA = osteoarthritis; TJR = total joint replacement; TKR = total knee replacement; EA = effect allele; beta = effect size (log(odds ratio)); pval = p-value*

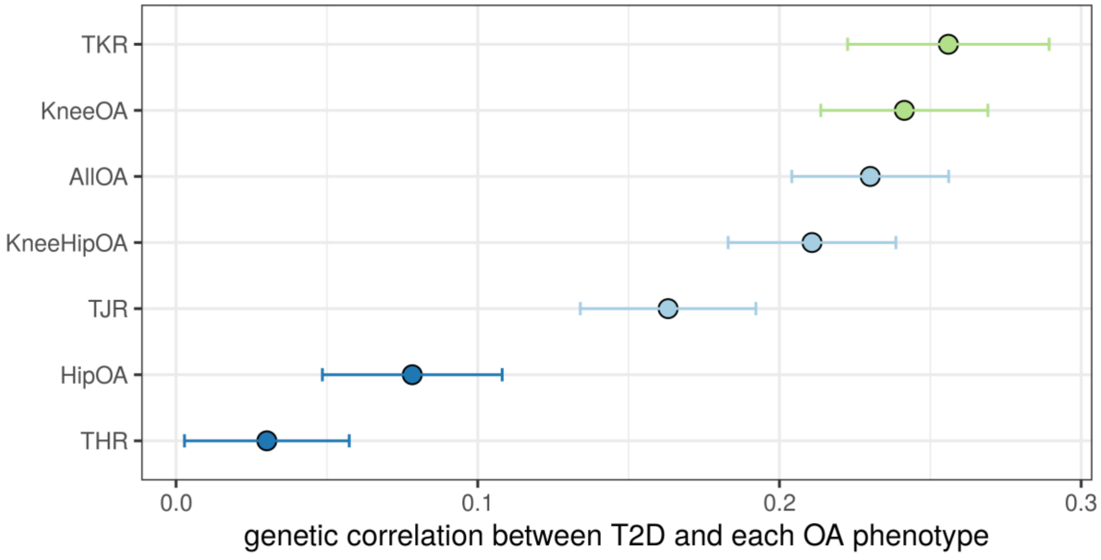


**Figure 1: Results of genetic correlation analysis between type 2 diabetes and osteoarthritis.** The y-axis depicts the osteoarthritis phenotype and the x-axis the respective genetic correlation. TKR = total knee replacement, THR = total hip replacement, AllOA = osteoarthritis at any site.

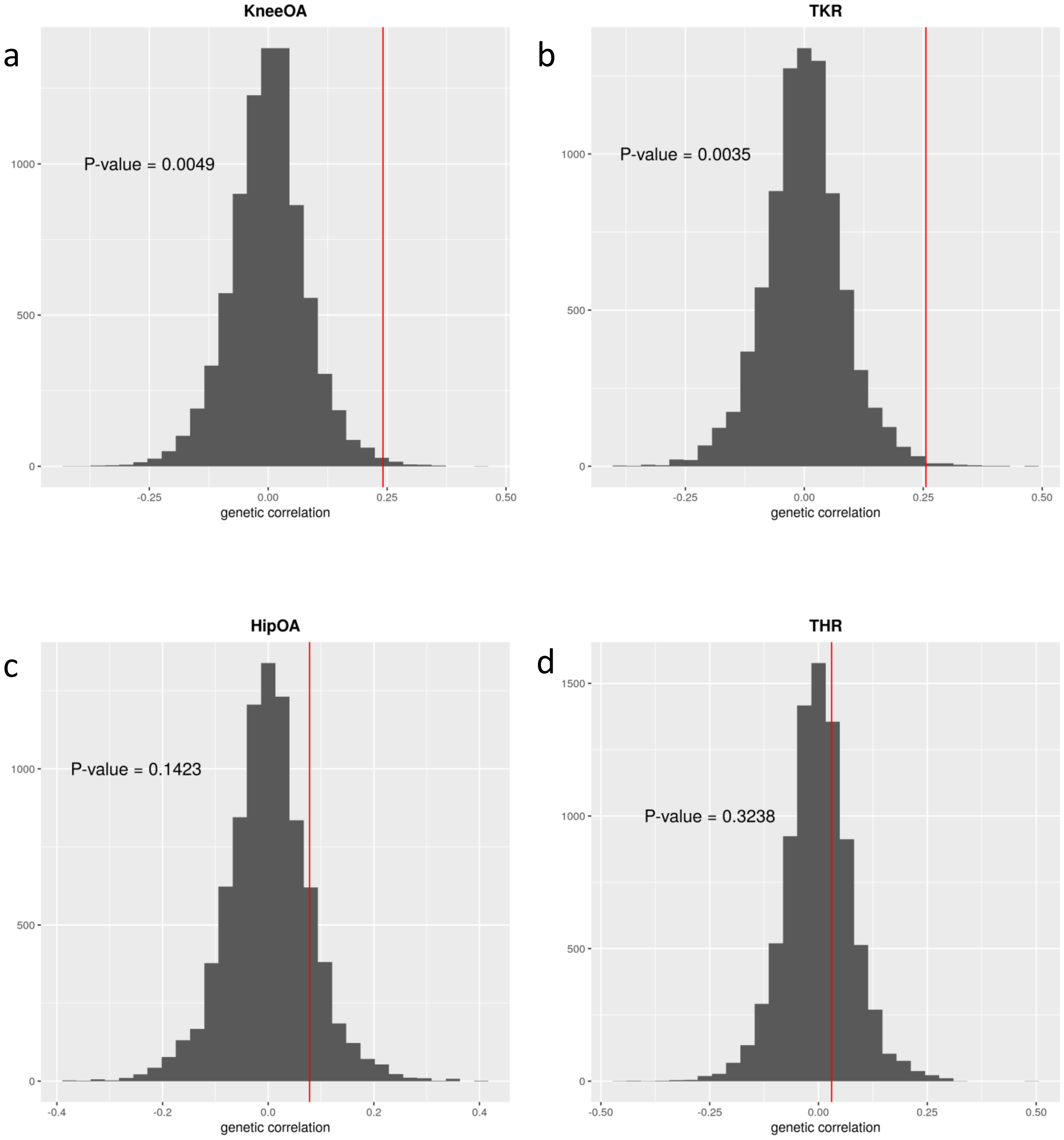
a
KneeOA
P-value = 0.0049
1000
500
0
-0.25
0.00
0.25
0.50
genetic correlation
b
TKR
P-value = 0.0035
1000
500
0
-0.25
0.00
0.25
0.50
genetic correlation
c
HipOA
P-value = 0.1423
1000
500
0
-0.4
-0.2
0.0
0.2
0.4
genetic correlation
d
THR
P-value = 0.3238
1500
1000
500
0
-0.50
-0.25
0.00
0.25
0.50
genetic correlation

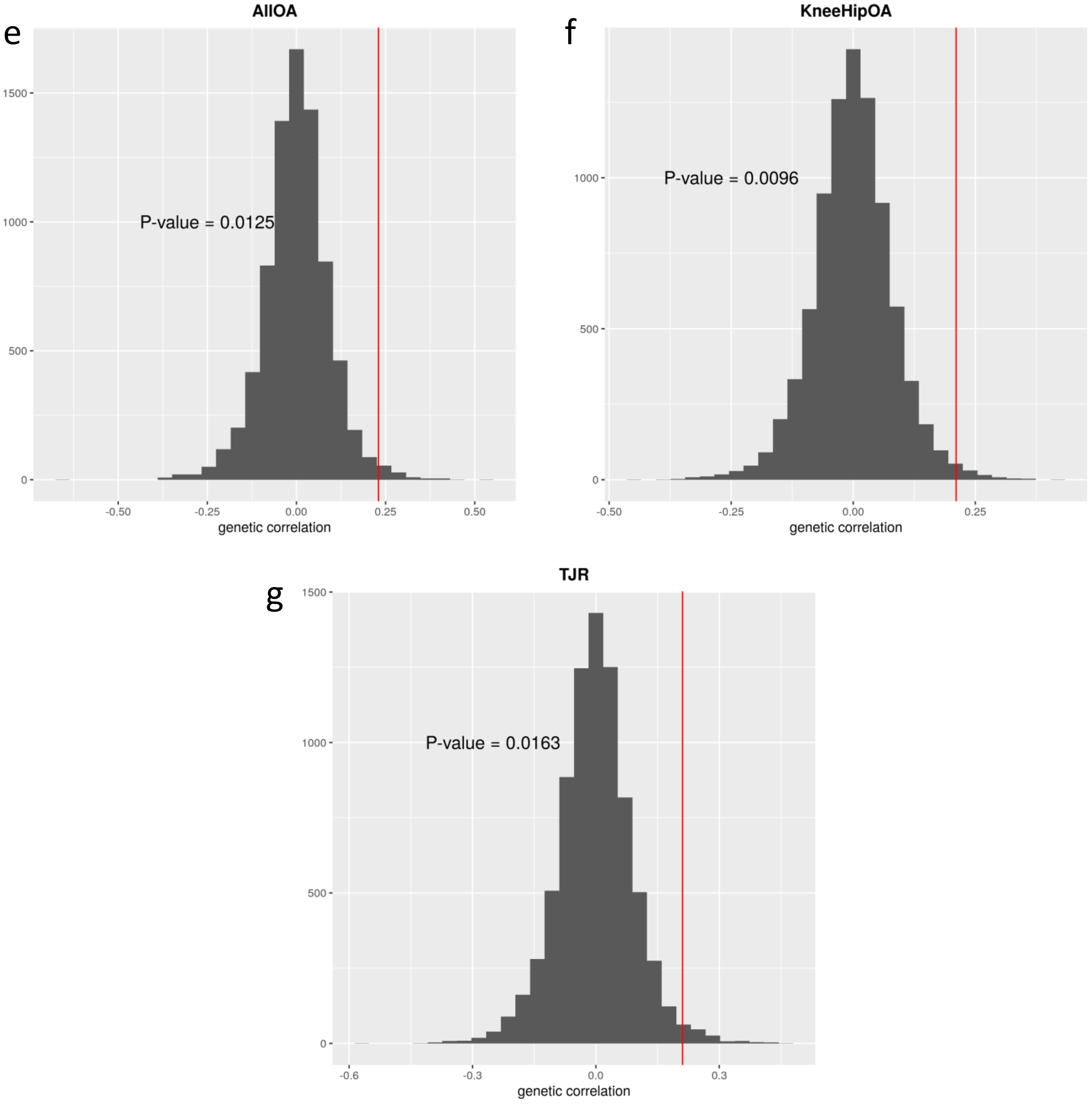


**Figure S2: Permutation-based testing of genetic correlation between type 2 diabetes and osteoarthritis.** The y-axis depicts the number of tests and the x-axis the genetic correlation of each test. The red line represents the true genetic correlation. Subfigures: a) genetic correlation between type 2 diabetes and knee osteoarthritis, b) genetic correlation between type 2 diabetes and total knee replacement (TKR), c) genetic correlation between type 2 diabetes and hip osteoarthritis, d) genetic correlation between type 2 diabetes and total hip replacement (THR), e) genetic correlation between type 2 diabetes and osteoarthritis at any site, f) genetic correlation between type 2 diabetes and knee and/or hip osteoarthritis, g) genetic correlation between type 2 diabetes and total joint replacement (TJR).

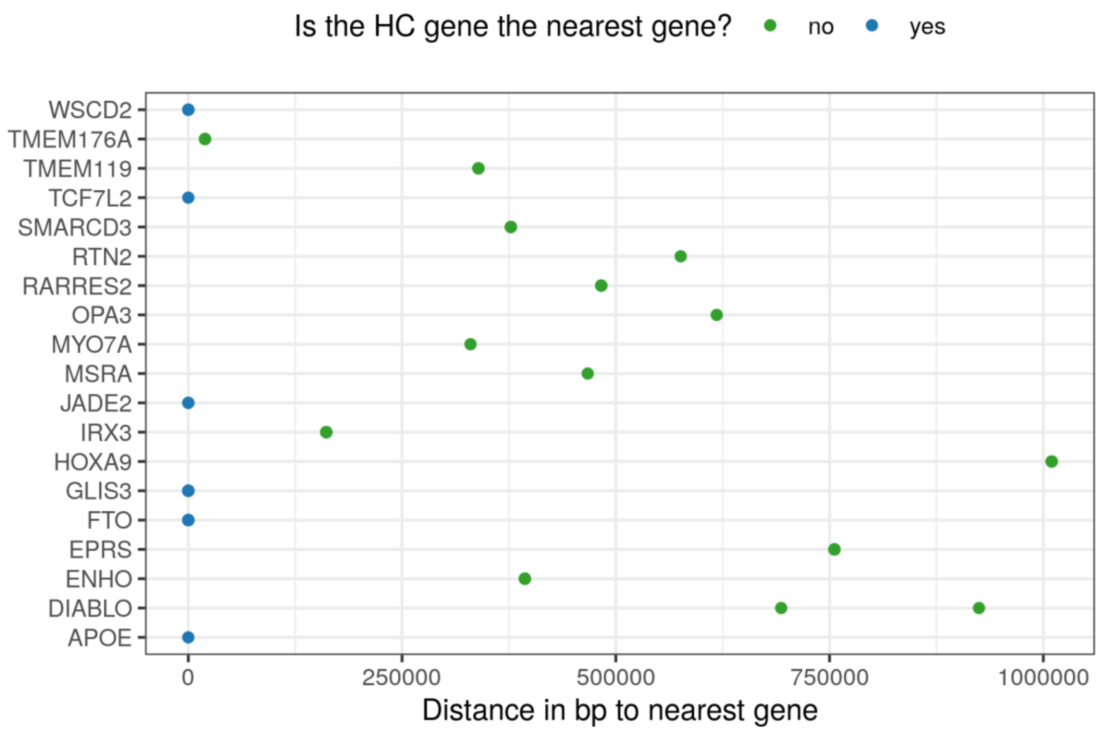


**Figure S3**: Distance in base pairs (bp) between each high confidence gene and the nearest gene to the lead causal variant in each colocalized region. Blue dots indicate that the high confidence gene is the nearest gene in the region.

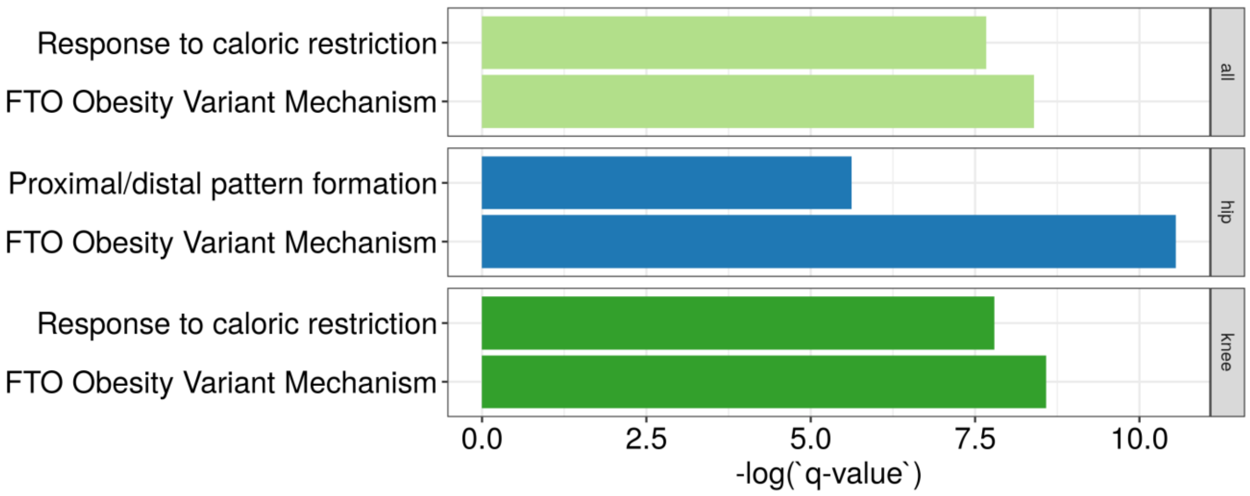


**Figure S4**: Top pathways enriched for the high confidence genes sets. The y-axis depicts the name of the enriched biological pathways for the different sets of genes: all = 19 high confidence genes, hip = 10 hip-related high confidence genes, knee = 18 knee-related high confidence genes. The x-axis depicts the negative logarithms of the FDR adjusted p-value (q-value).

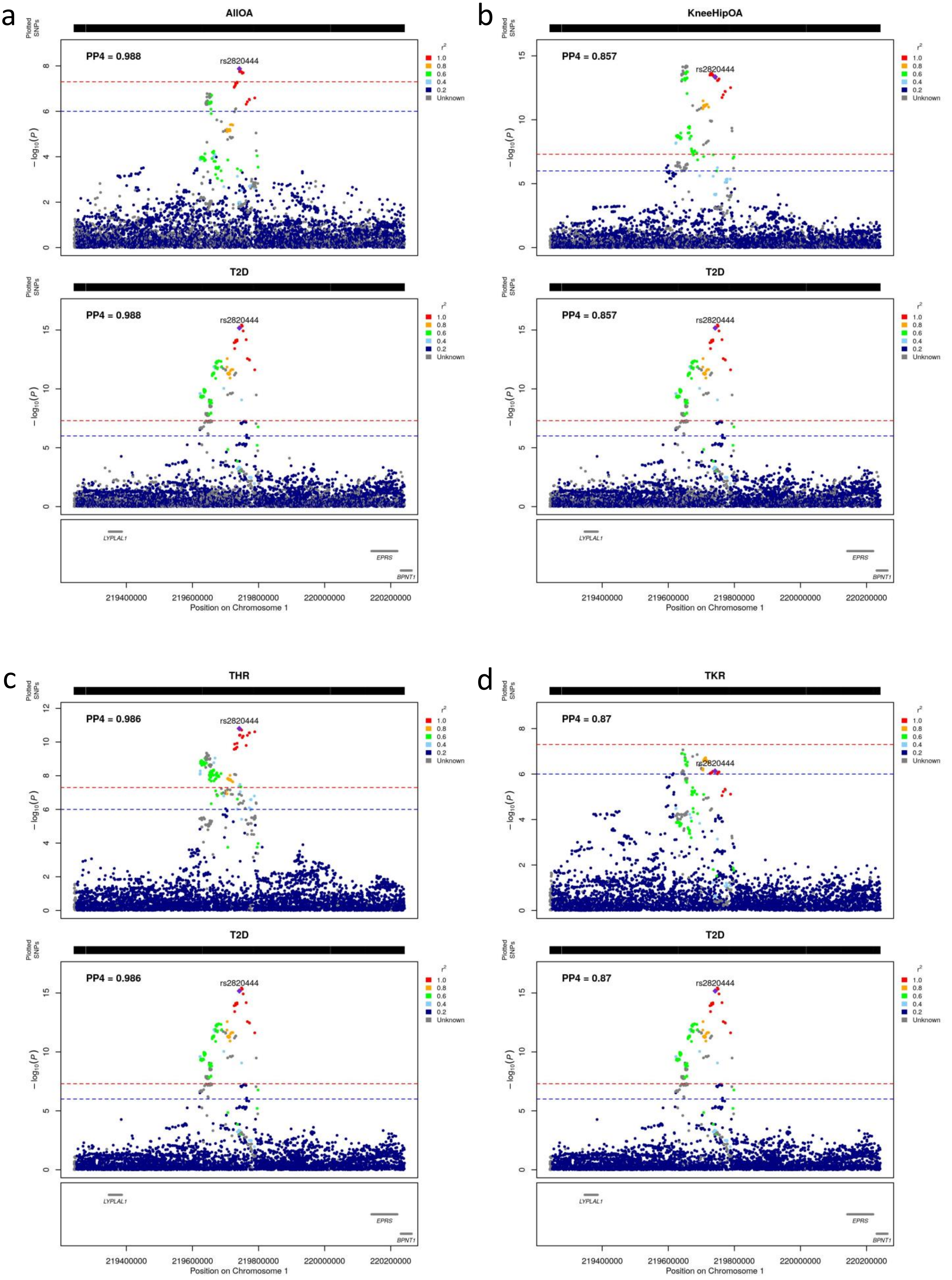
a
AllOA
PP4 = 0.988
rs2820444
T2D
PP4 = 0.988
rs2820444
b
KneeHipOA
PP4 = 0.857
rs2820444
T2D
PP4 = 0.857
rs2820444
c
THR
PP4 = 0.986
rs2820444
T2D
PP4 = 0.986
rs2820444
d
TKR
PP4 = 0.87
rs2820444
T2D
PP4 = 0.87
rs2820444
Plotted SNPs
$-\log_{10}(P)$
$r^2$
1.0
0.8
0.6
0.4
0.2
Unknown
LYPLAL1
EPRS
BPNT1
219400000
219600000
219800000
220000000
220200000
Position on Chromosome 1

e

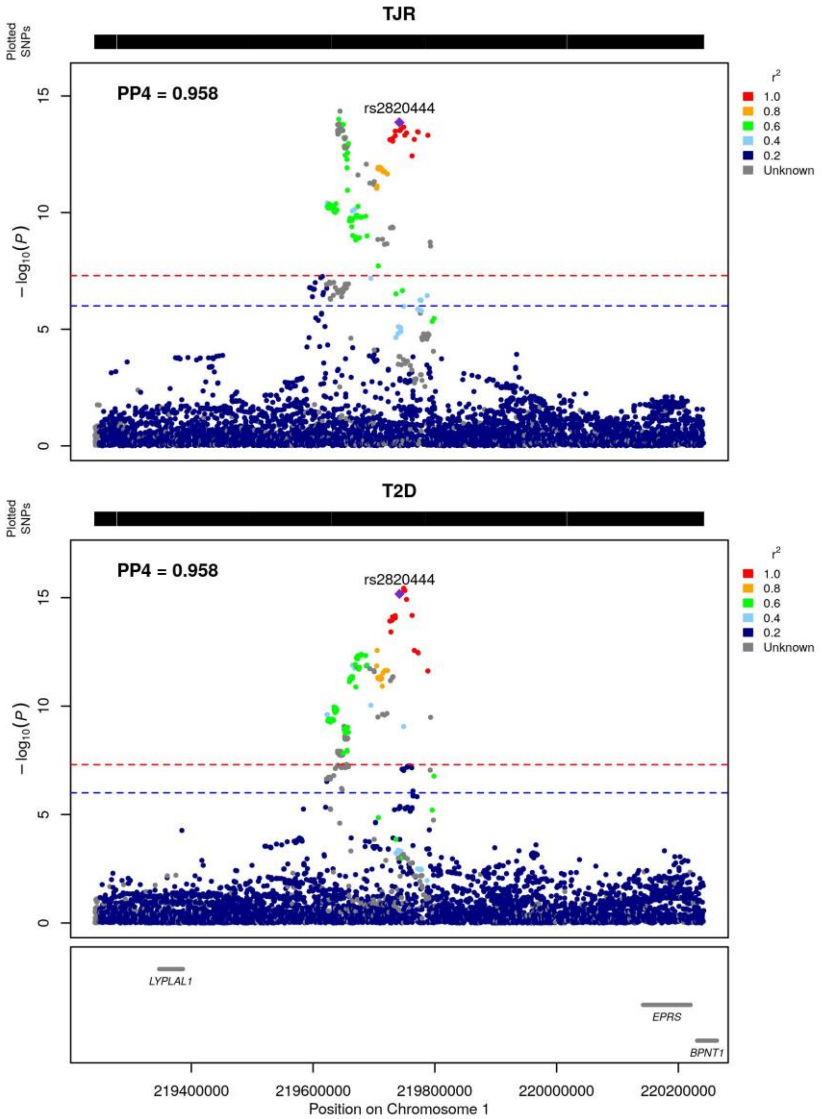


**Figure S5: Colocalized region 1.** This region colocalizes between type 2 diabetes and a) osteoarthritis at any site , b) knee and/or hip osteoarthritis, c) total hip replacement, d) total knee replacement, e) total joint replacement.

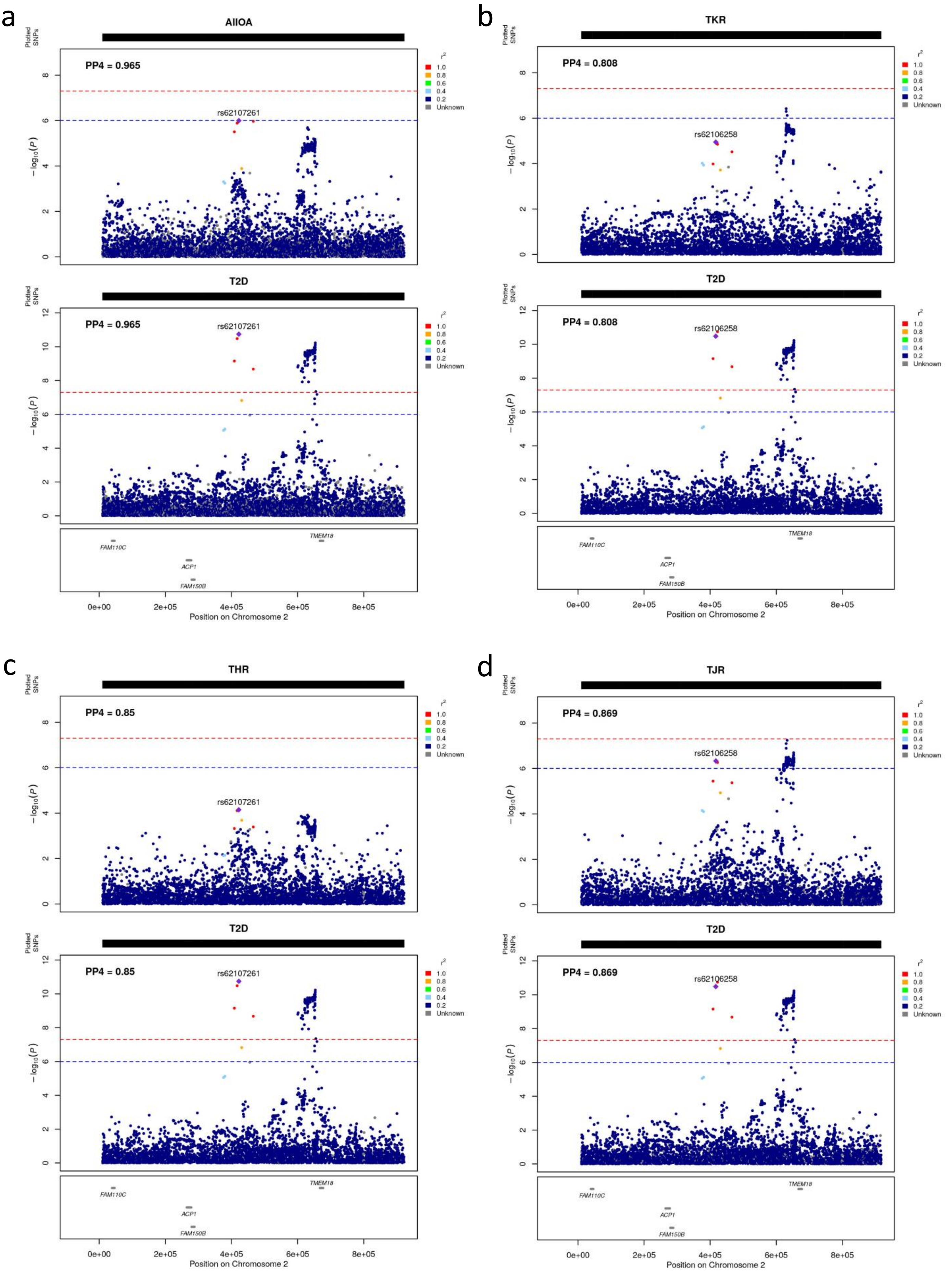


**Figure S6: Colocalized region 2.** This region colocalizes between type 2 diabetes and a) osteoarthritis at any site , b) total knee replacement, c) total hip replacement, d) total joint replacement.

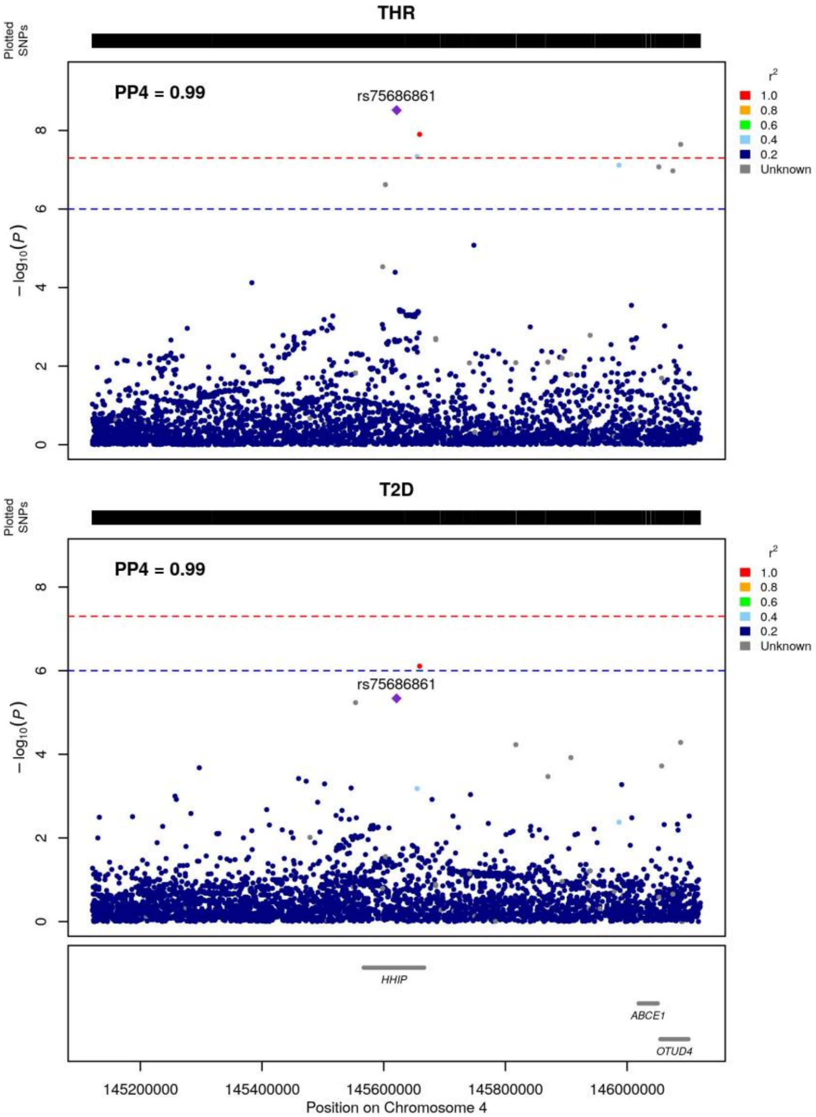


**Figure S7: Colocalized region 3.** This region colocalizes between type 2 diabetes and total hip replacement.

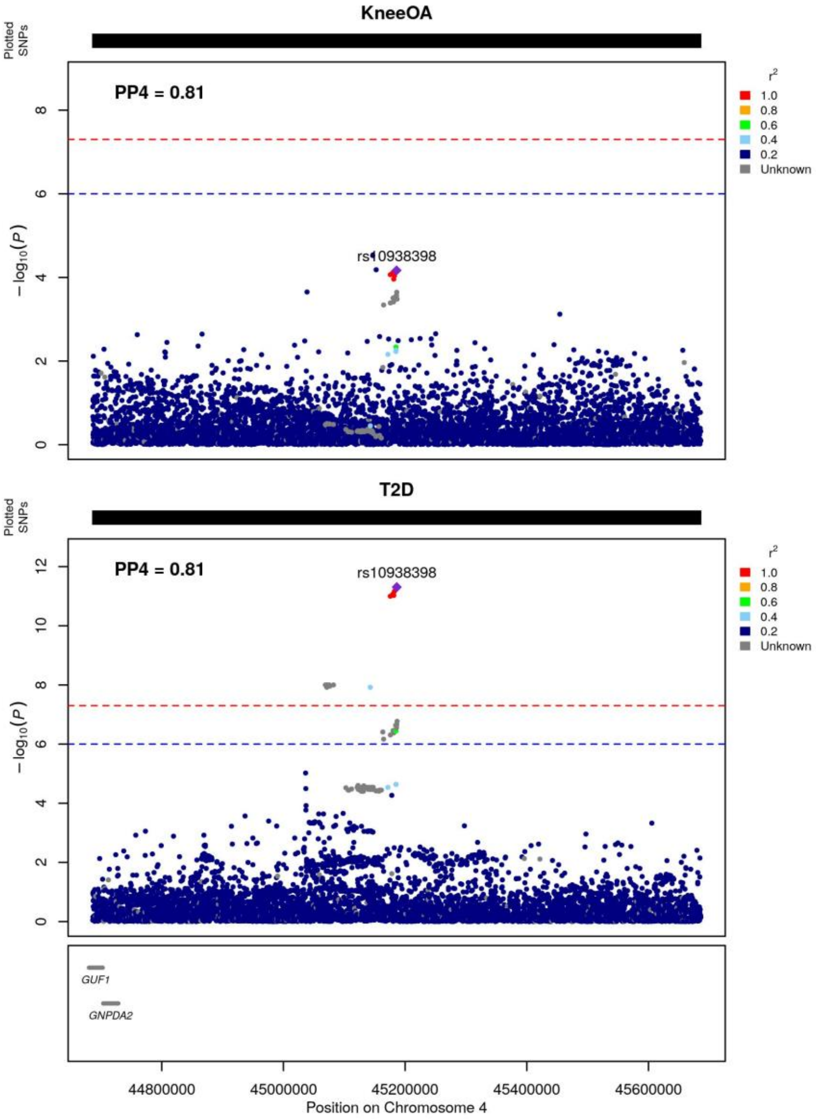


**Figure S8: Colocalized region 4.** This region colocalizes between type 2 diabetes and knee osteoarthritis.

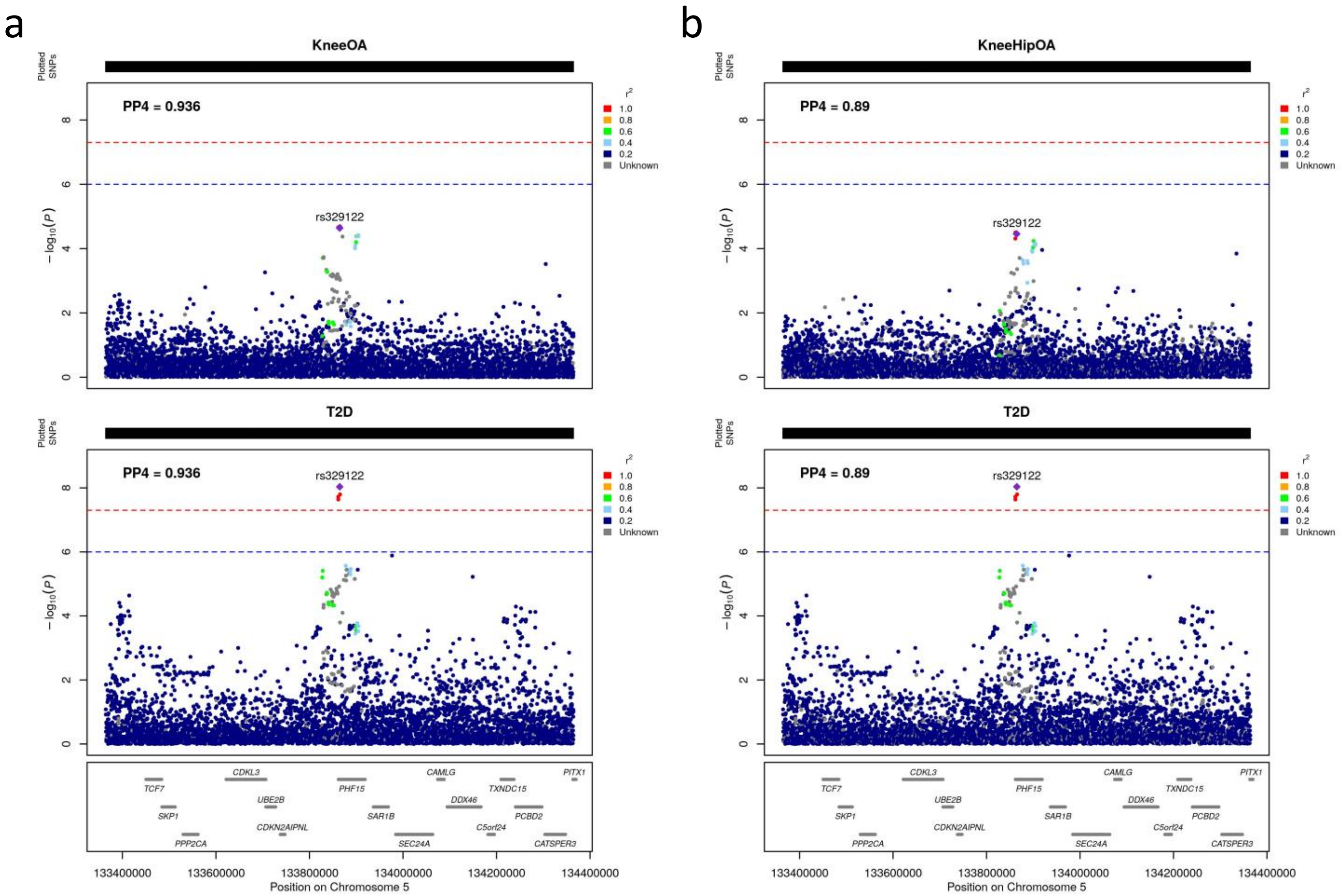


**Figure S9: Colocalized region 5.** This region colocalizes between type 2 diabetes and a) knee osteoarthritis, b) knee and/or hip osteoarthritis.

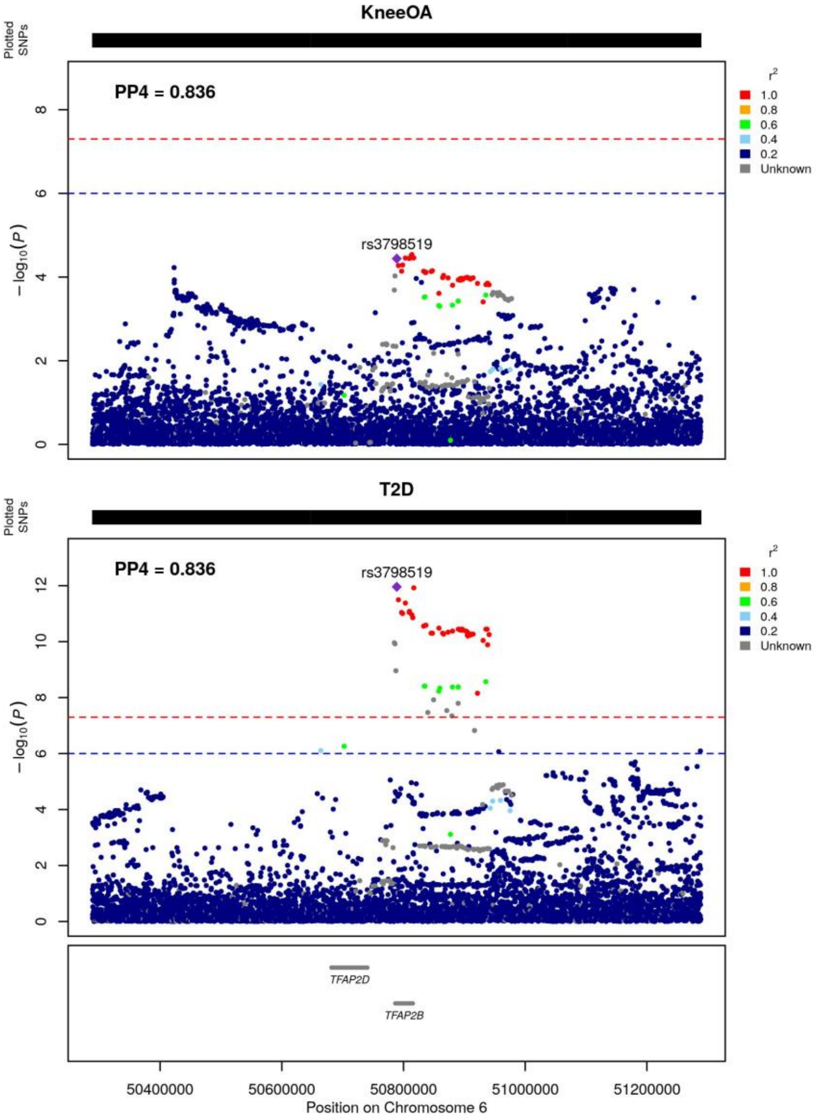


**Figure S10: Colocalized region 6.** This region colocalizes between type 2 diabetes and knee osteoarthritis.

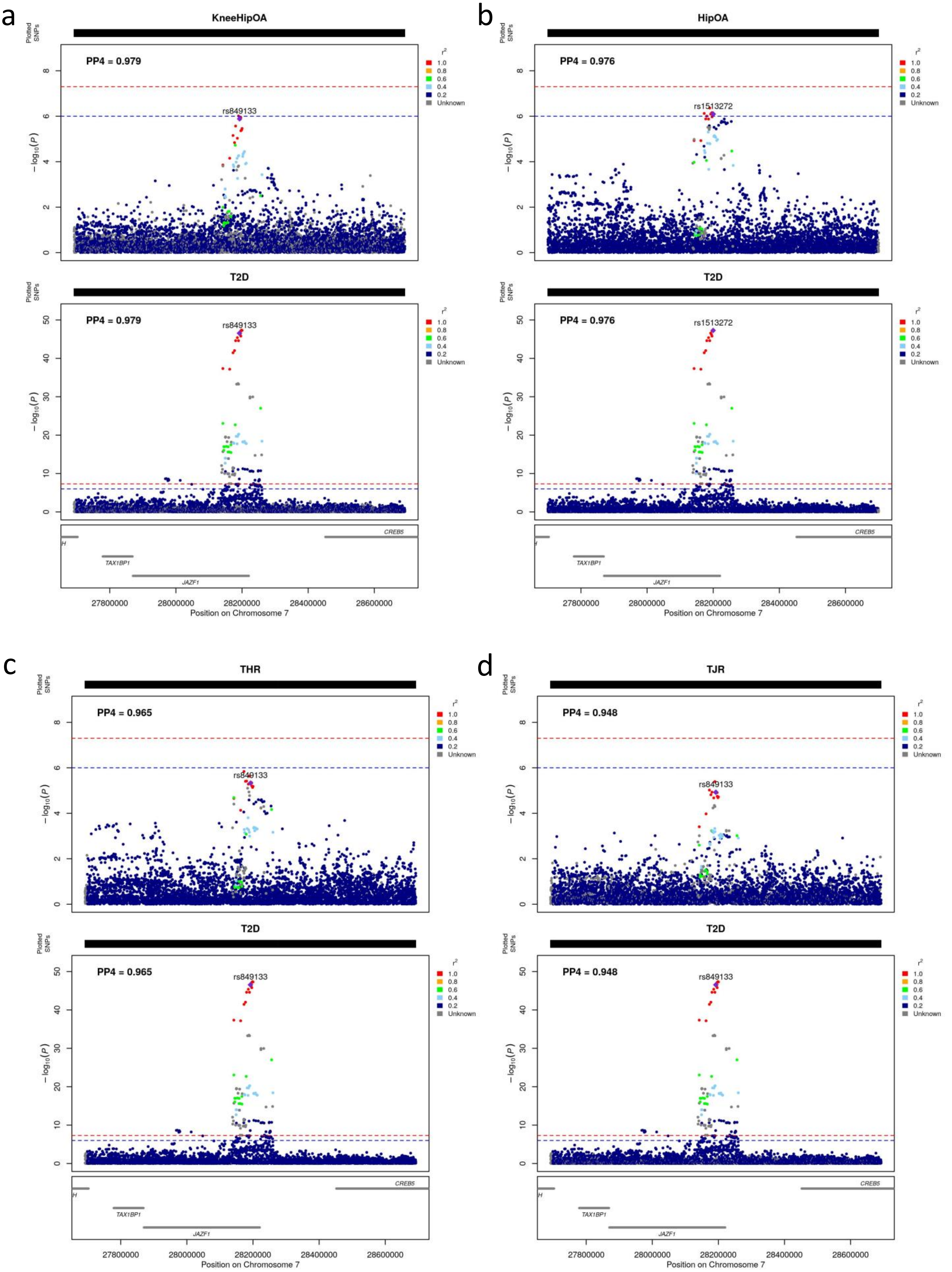


**Figure S11: Colocalized region 7.** This region colocalizes between type 2 diabetes and a) knee and/or hip osteoarthritis, b) hip osteoarthritis, c) total hip replacement, d) total joint replacement.

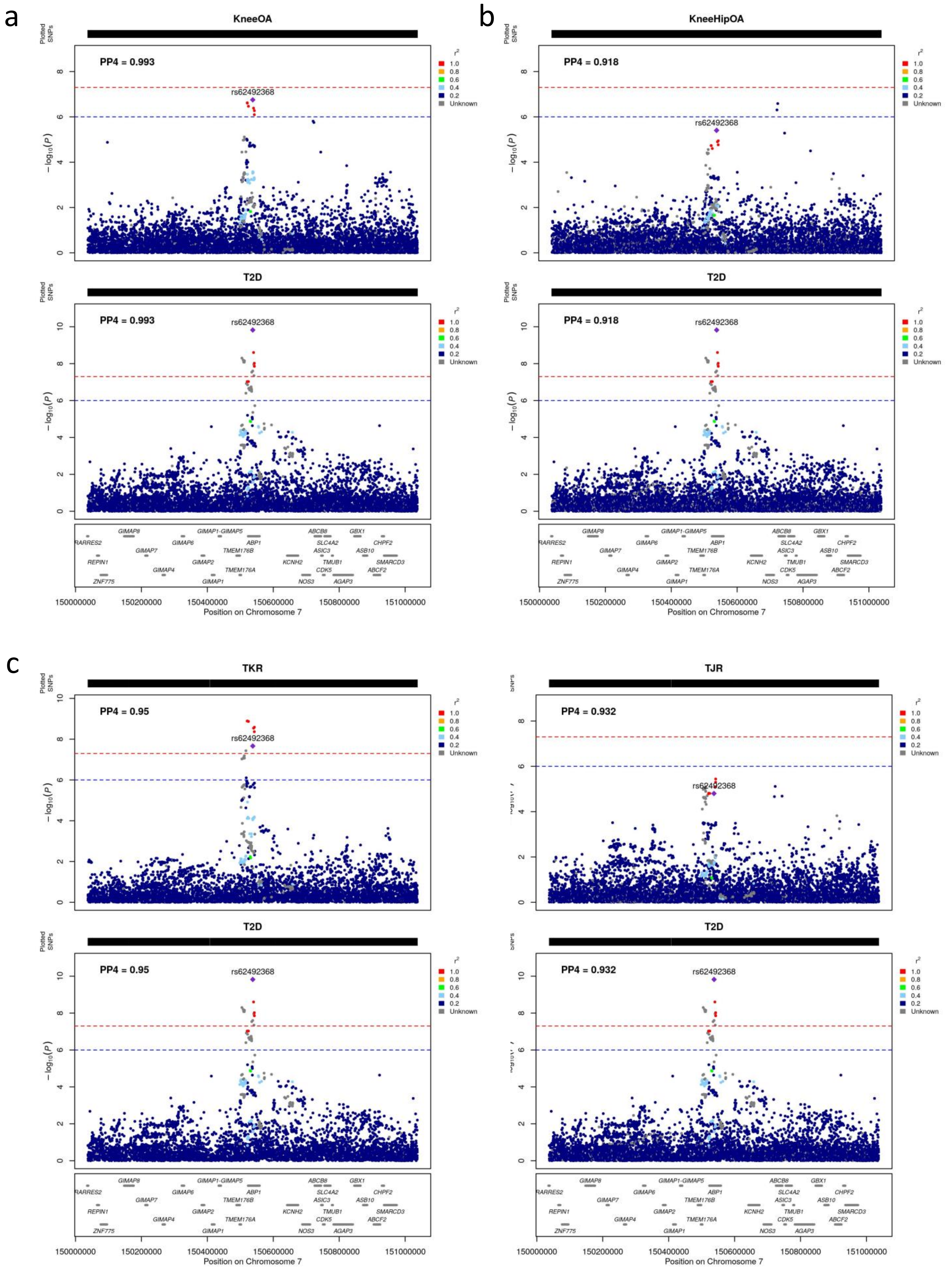


**Figure S12: Colocalized region 8.** This region colocalizes between type 2 diabetes and a) knee osteoarthritis, b) knee and/or hip osteoarthritis, c) total knee replacement, d) total joint replacement.

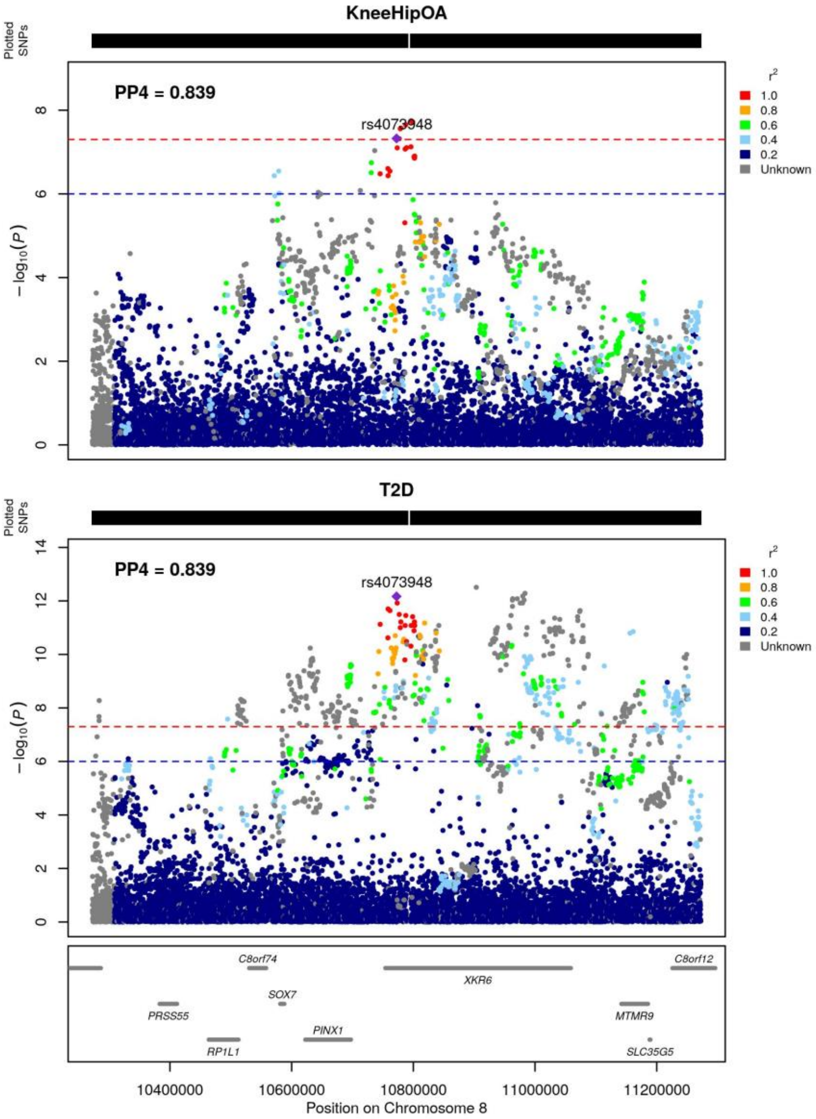


**Figure S13: Colocalized region 9.** This region colocalizes between type 2 diabetes and knee and/or hip osteoarthritis.

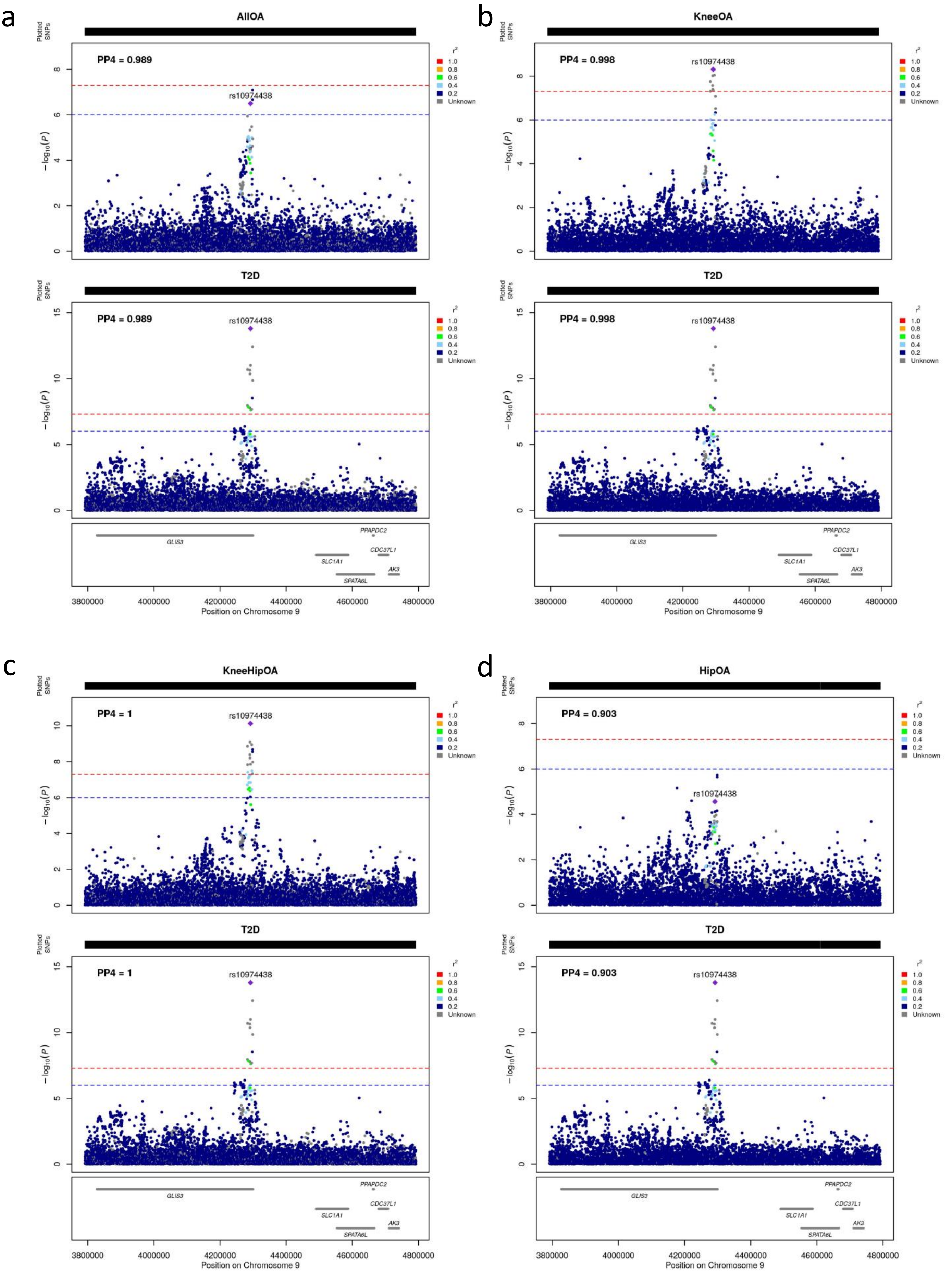
a
AllOA
PP4 = 0.989
rs10974438
T2D
PP4 = 0.989
rs10974438
b
KneeOA
PP4 = 0.998
rs10974438
T2D
PP4 = 0.998
rs10974438
c
KneeHipOA
PP4 = 1
rs10974438
T2D
PP4 = 1
rs10974438
d
HipOA
PP4 = 0.903
rs10974438
T2D
PP4 = 0.903
rs10974438
Plotted SNPs
$-\log_{10}(P)$
$r^2$
1.0
0.8
0.6
0.4
0.2
Unknown
GLIS3
PPAPDC2
CDC37L1
SLC1A1
AK3
SPATA6L
3800000
4000000
4200000
4400000
4600000
4800000
Position on Chromosome 9

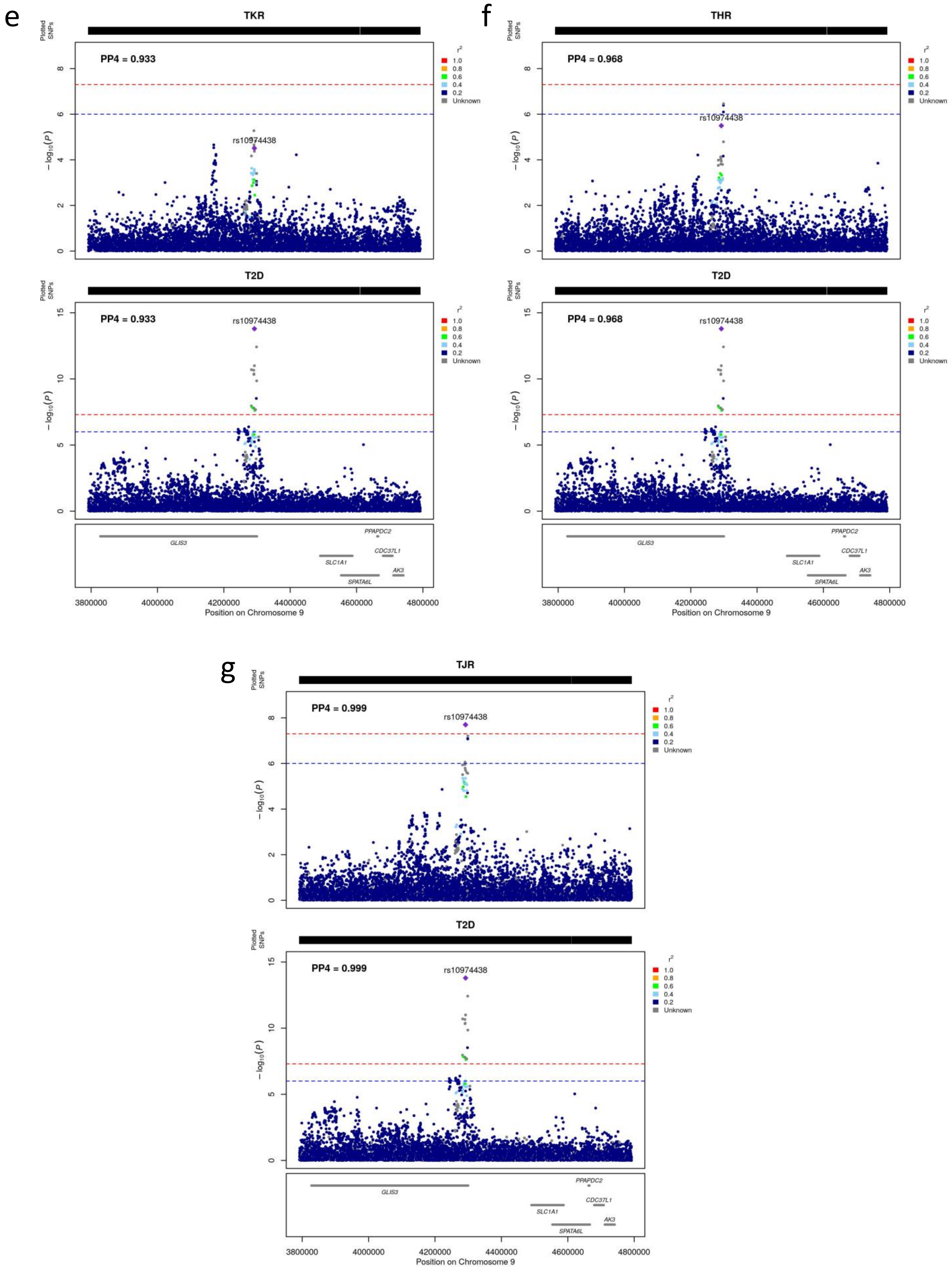


**Figure S14: Colocalized region 10.** This region colocalizes between type 2 diabetes and a) osteoarthritis at any site, b) knee osteoarthritis, c) knee and/or hip osteoarthritis, d) hip osteoarthritis, e) total knee replacement, f) total hip replacement, g) total joint replacement.

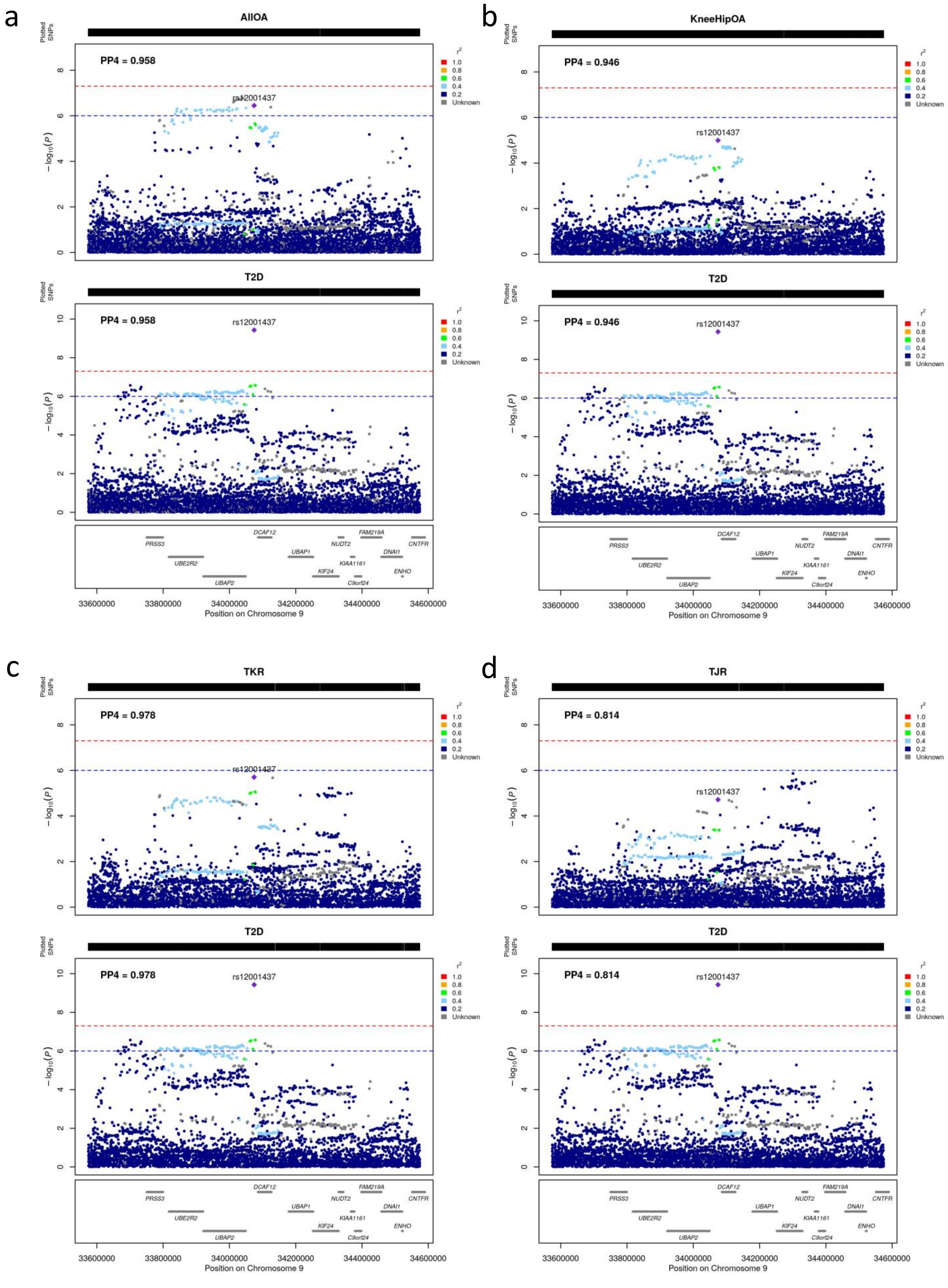


**Figure S15: Colocalized region 11.** This region colocalizes between type 2 diabetes and a) osteoarthritis at any site, b) knee and/or hip osteoarthritis, c) total knee replacement, d) total joint replacement.

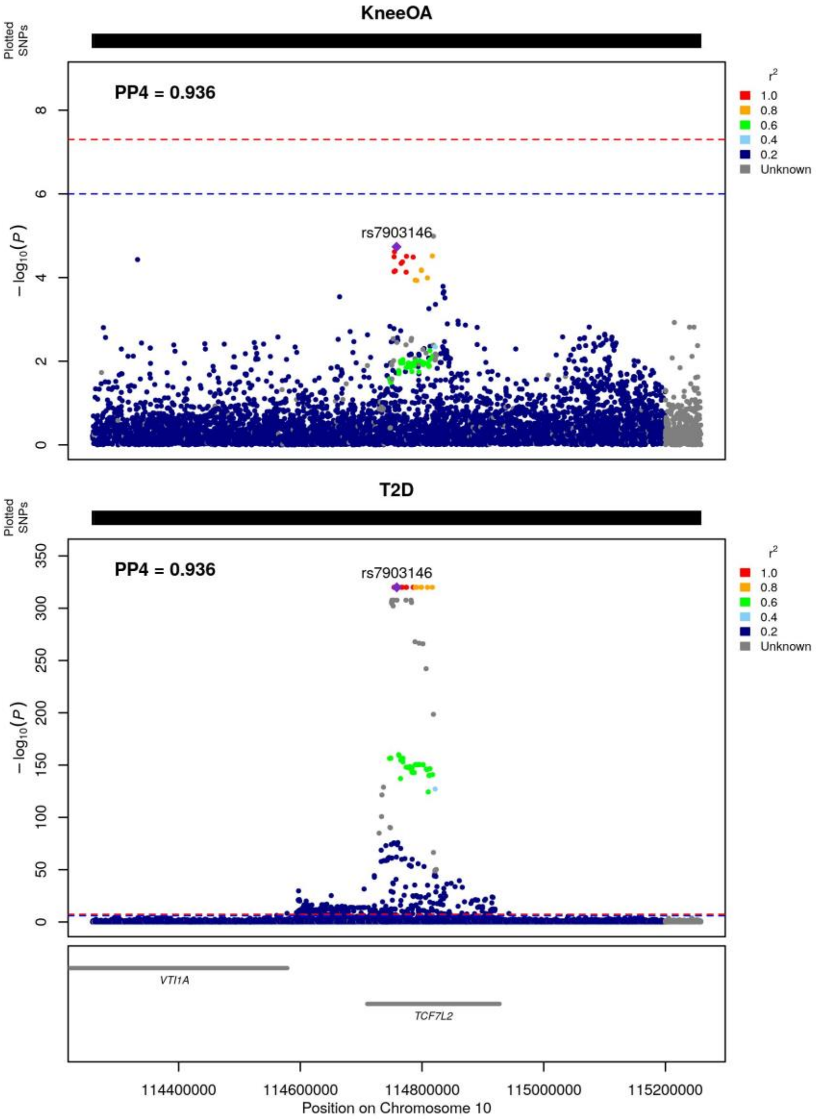


**Figure S16: Colocalized region 12.** This region colocalizes between type 2 diabetes and knee osteoarthritis.

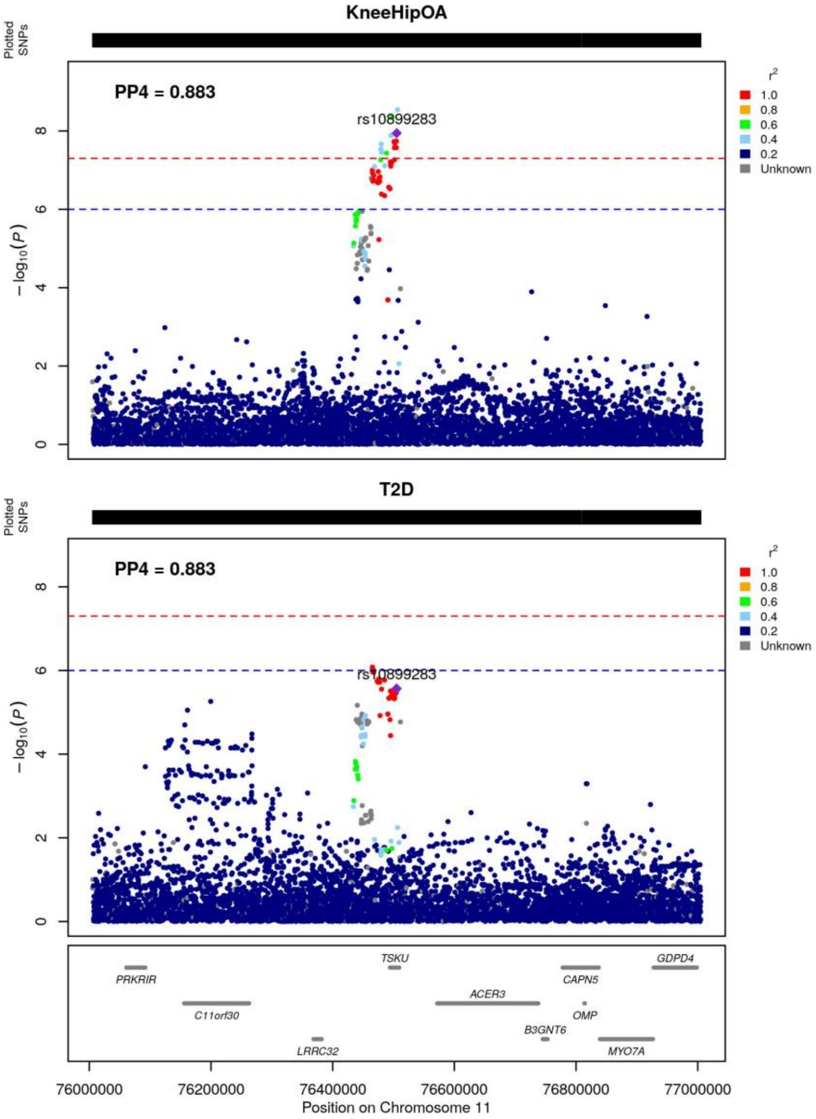


**Figure S17: Colocalized region 13.** This region colocalizes between type 2 diabetes and knee and/or hip osteoarthritis.

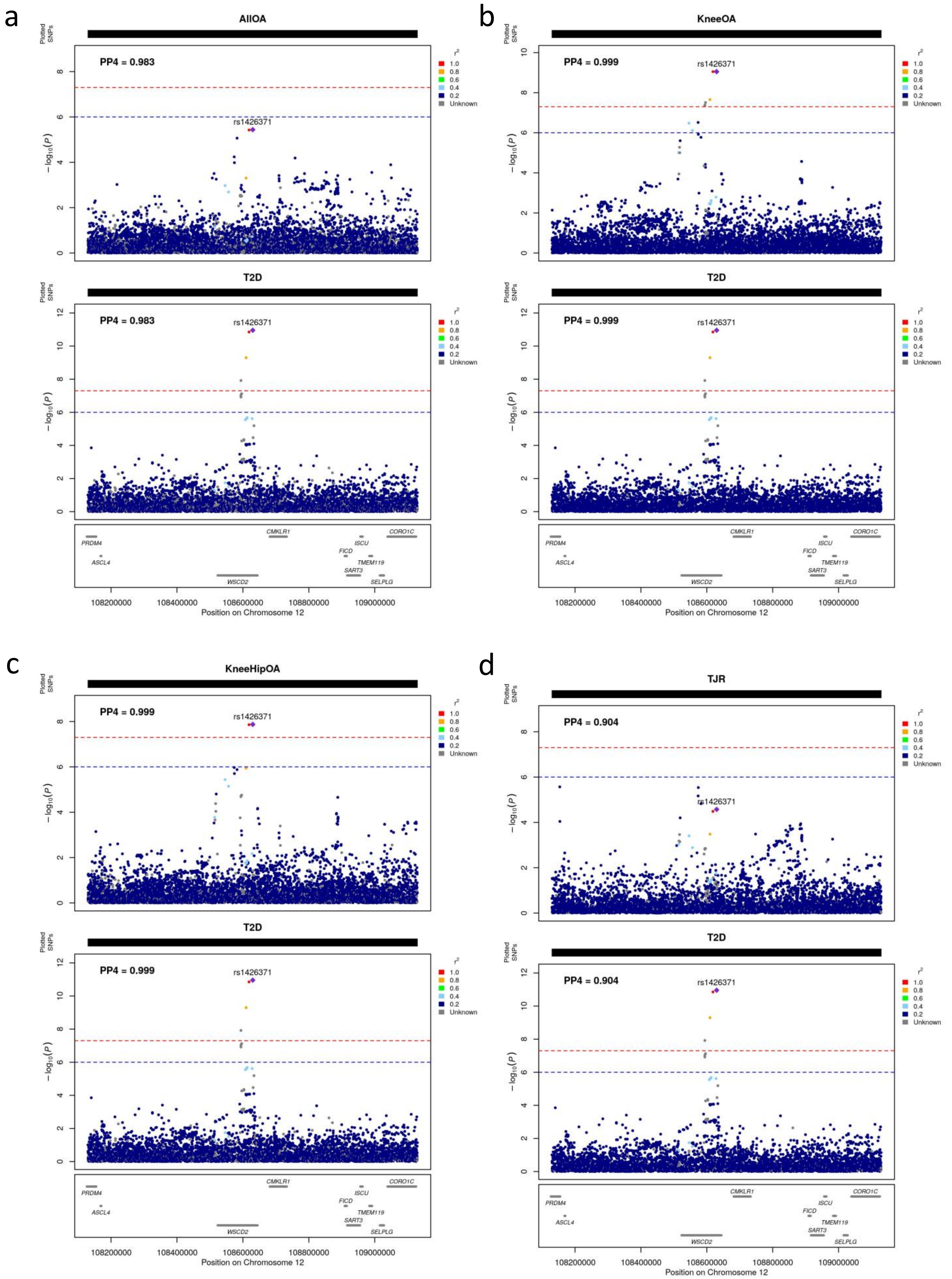


**Figure S18: Colocalized region 14.** This region colocalizes between type 2 diabetes and a) osteoarthritis at any site, b) knee osteoarthritis, c) knee and/or hip osteoarthritis, d) total joint replacement.

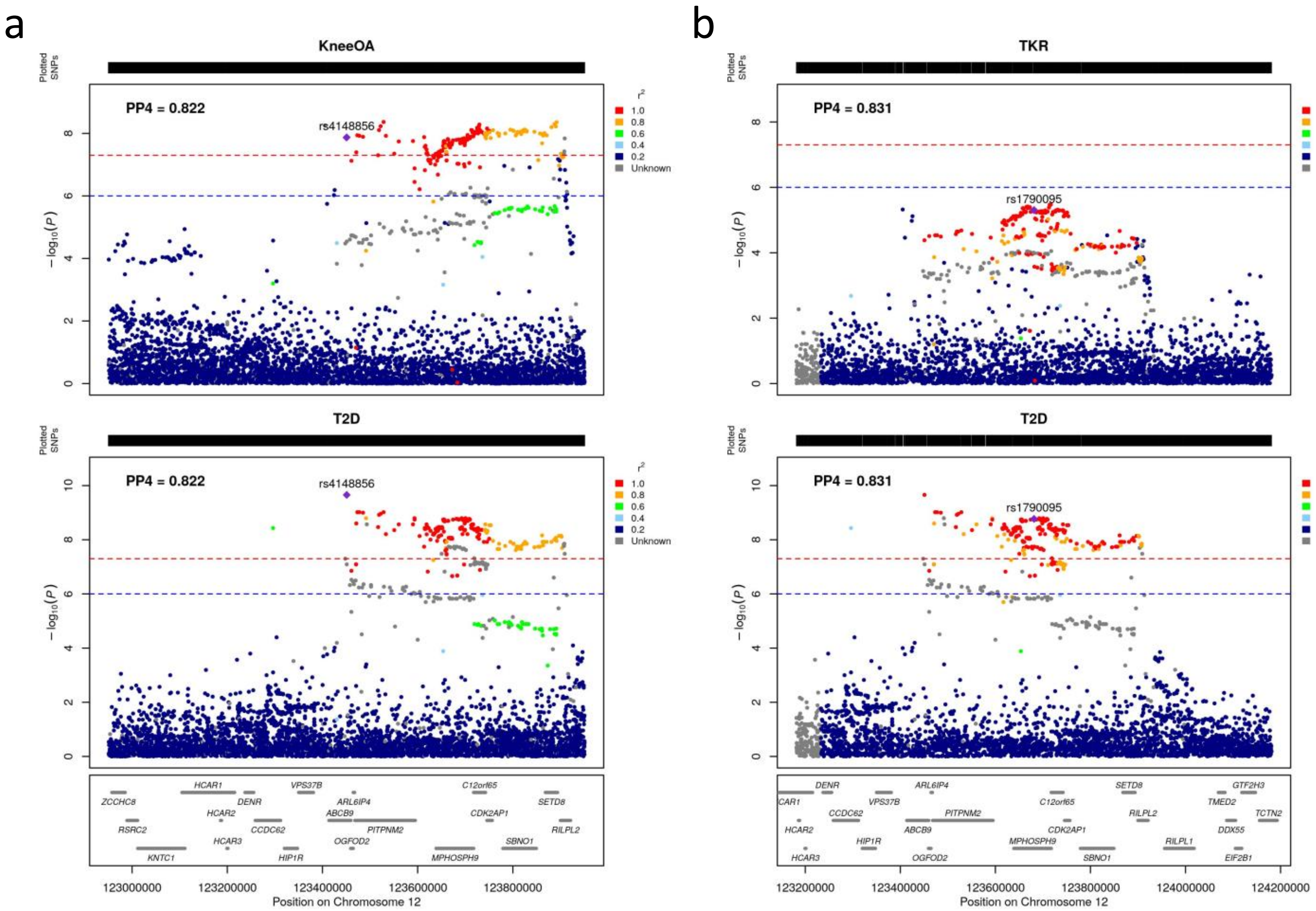


**Figure S19: Colocalized region 15.** This region colocalizes between type 2 diabetes and a) knee osteoarthritis, b) total knee replacement.

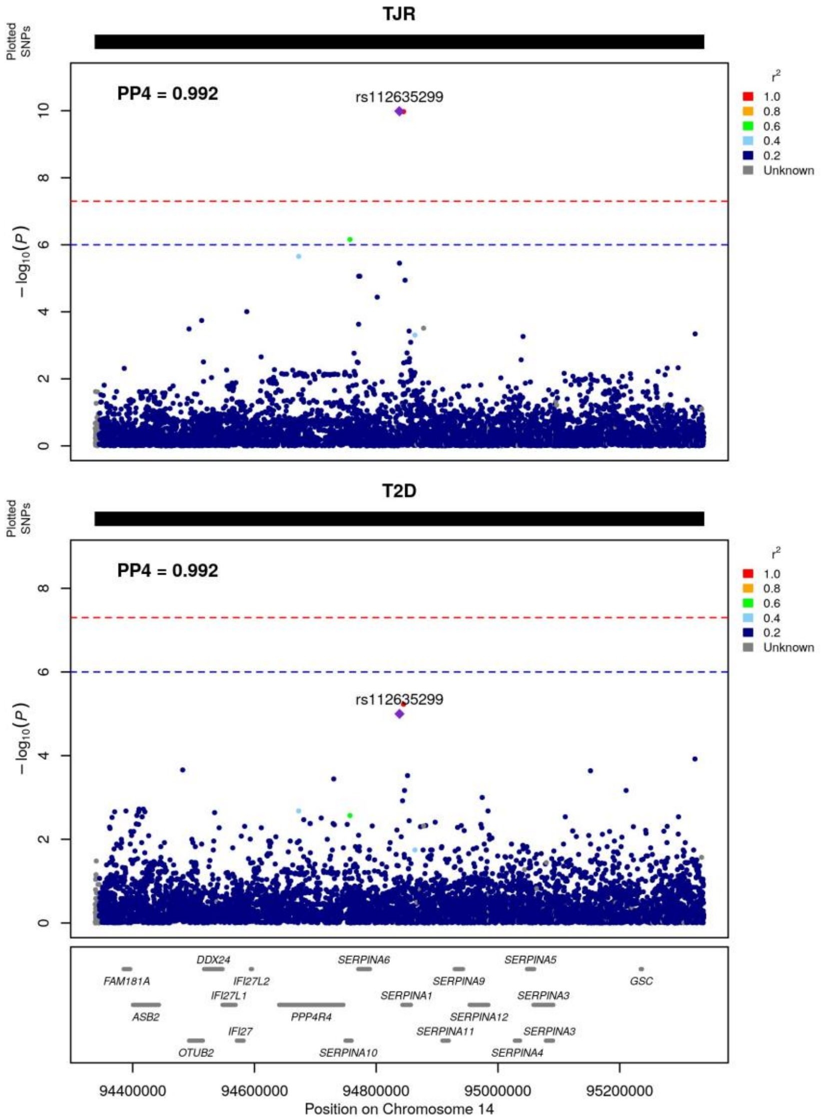


**Figure S20: Colocalized region 16.** This region colocalizes between type 2 diabetes and total joint replacement.

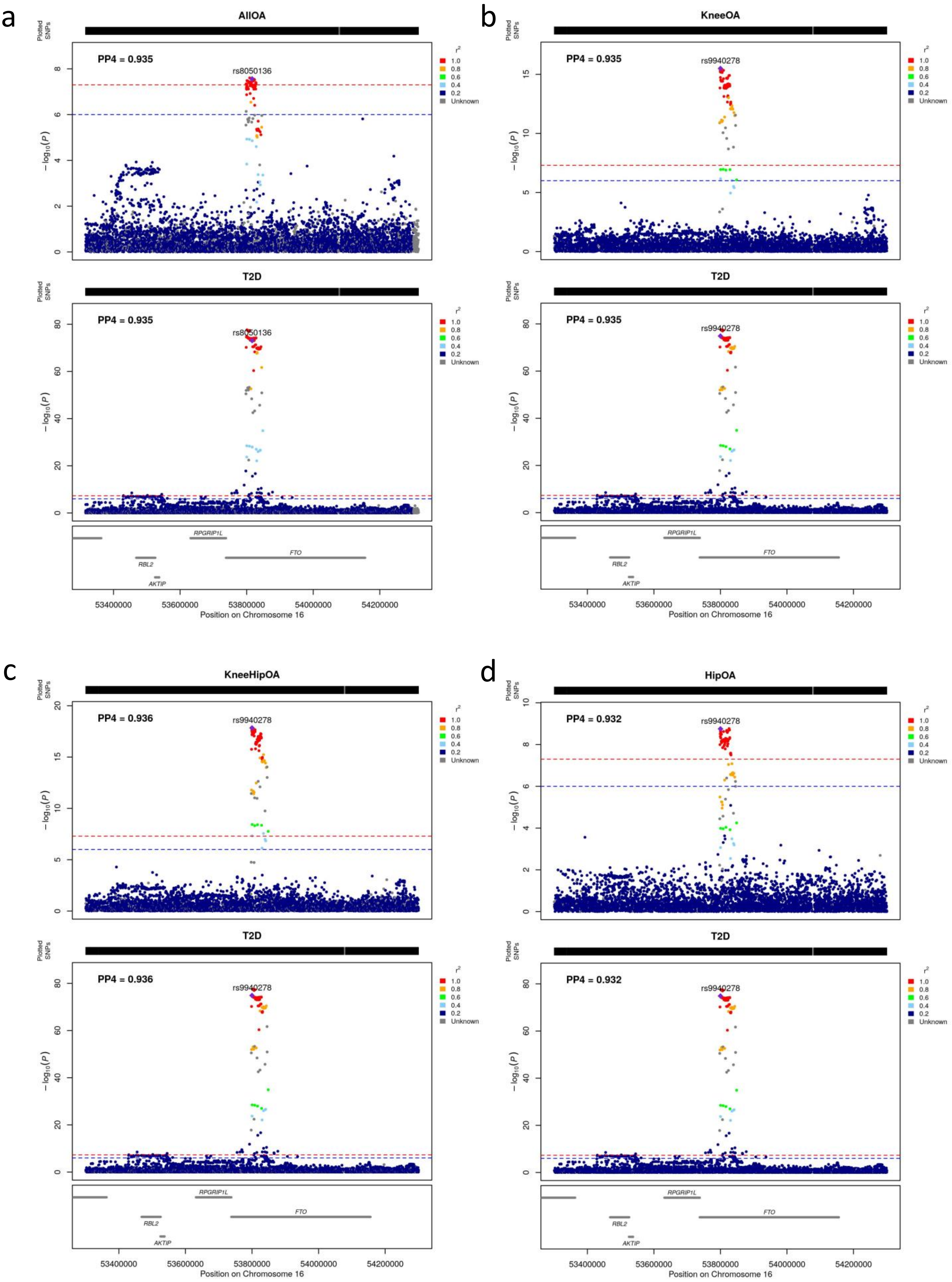
a
AllOA
Plotted SNPs
PP4 = 0.935
rs8050136
r²
1.0
0.8
0.6
0.4
0.2
Unknown
−log₁₀(P)
T2D
PP4 = 0.935
rs8050136
RPGRIP1L
FTO
RBL2
AKTIP
53400000
53600000
53800000
54000000
54200000
Position on Chromosome 16
b
KneeOA
PP4 = 0.935
rs9940278
T2D
PP4 = 0.935
rs9940278
Position on Chromosome 16
c
KneeHipOA
PP4 = 0.936
rs9940278
T2D
PP4 = 0.936
rs9940278
Position on Chromosome 16
d
HipOA
PP4 = 0.932
rs9940278
T2D
PP4 = 0.932
rs9940278
Position on Chromosome 16

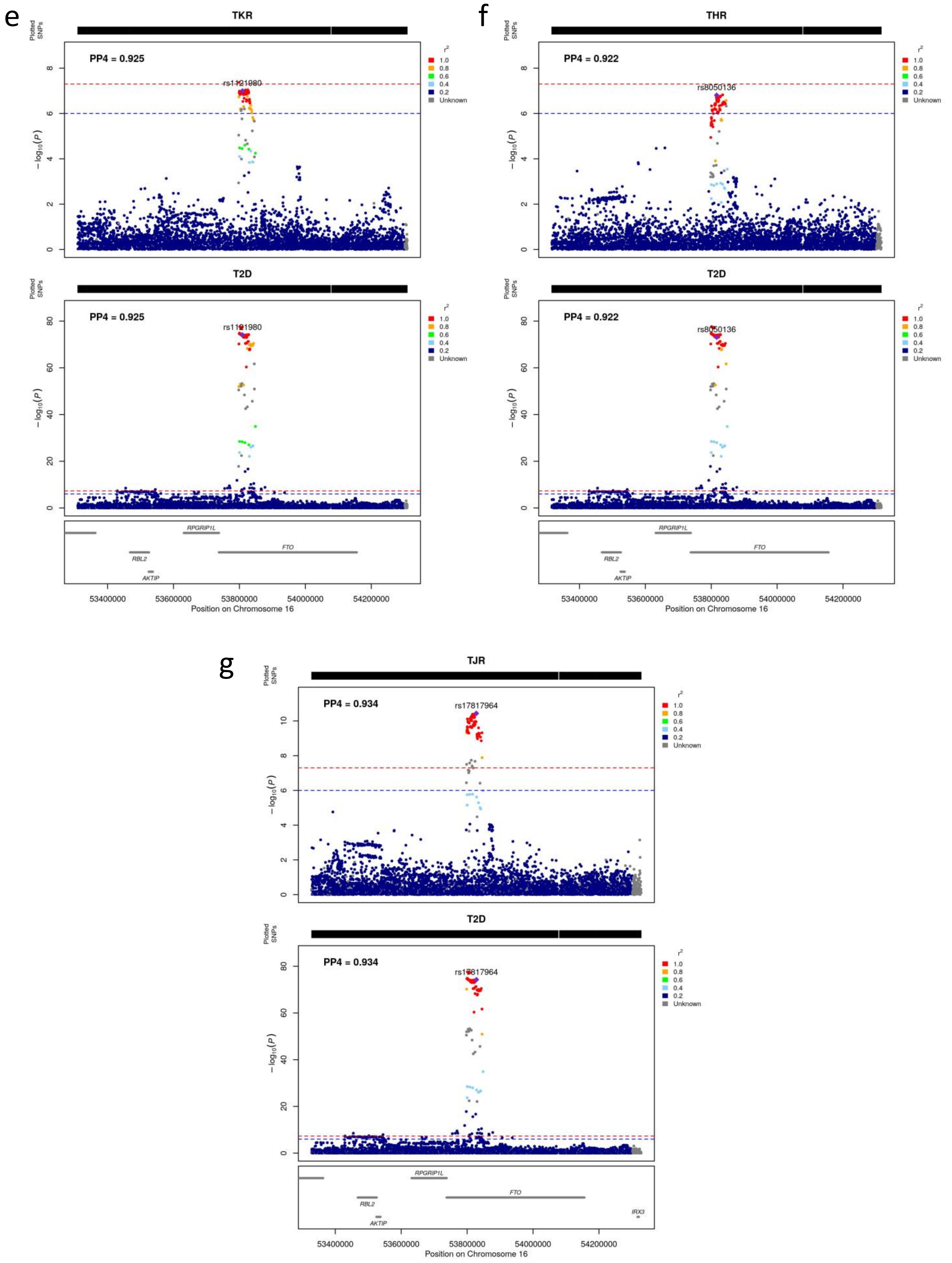


**Figure S21: Colocalized region 17.** This region colocalizes between type 2 diabetes and a) osteoarthritis at any site, b) knee osteoarthritis, c) knee and/or hip osteoarthritis, d) hip osteoarthritis, e) total knee replacement, f) total hip replacement, g) total joint replacement.

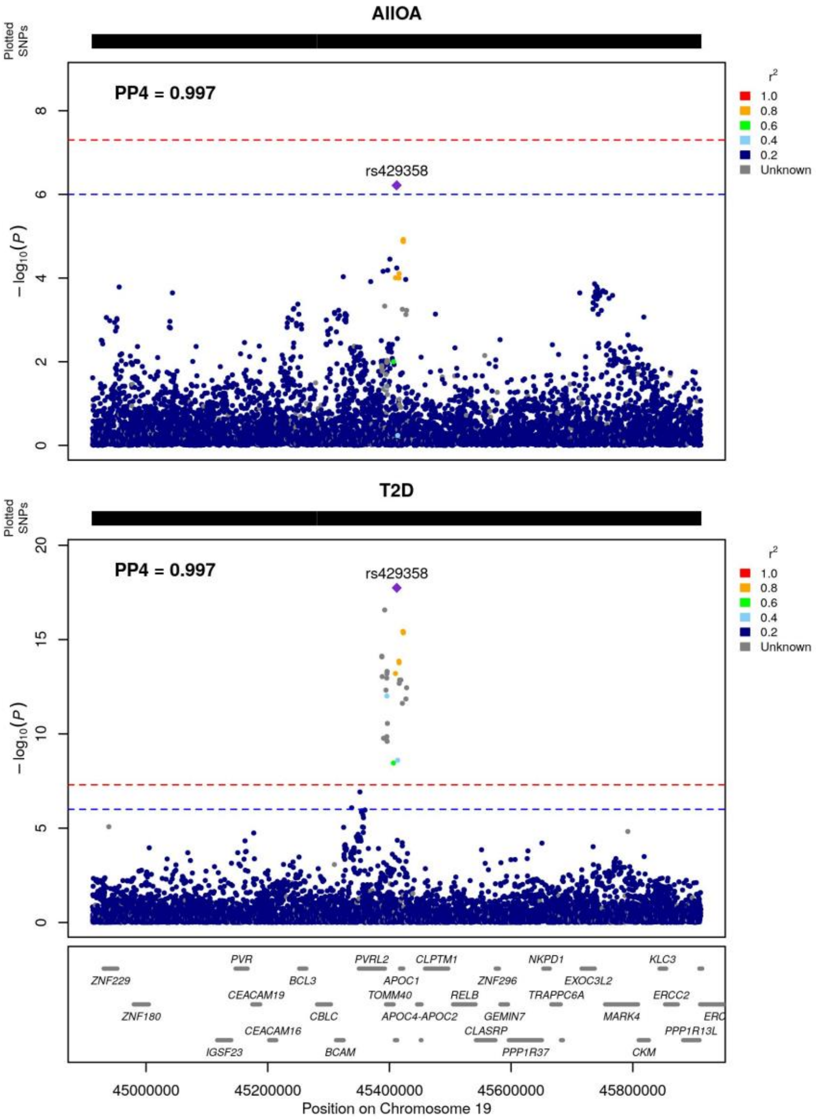


**Figure S22: Colocalized region 18.** This region colocalizes between type 2 diabetes and osteoarthritis at any site.

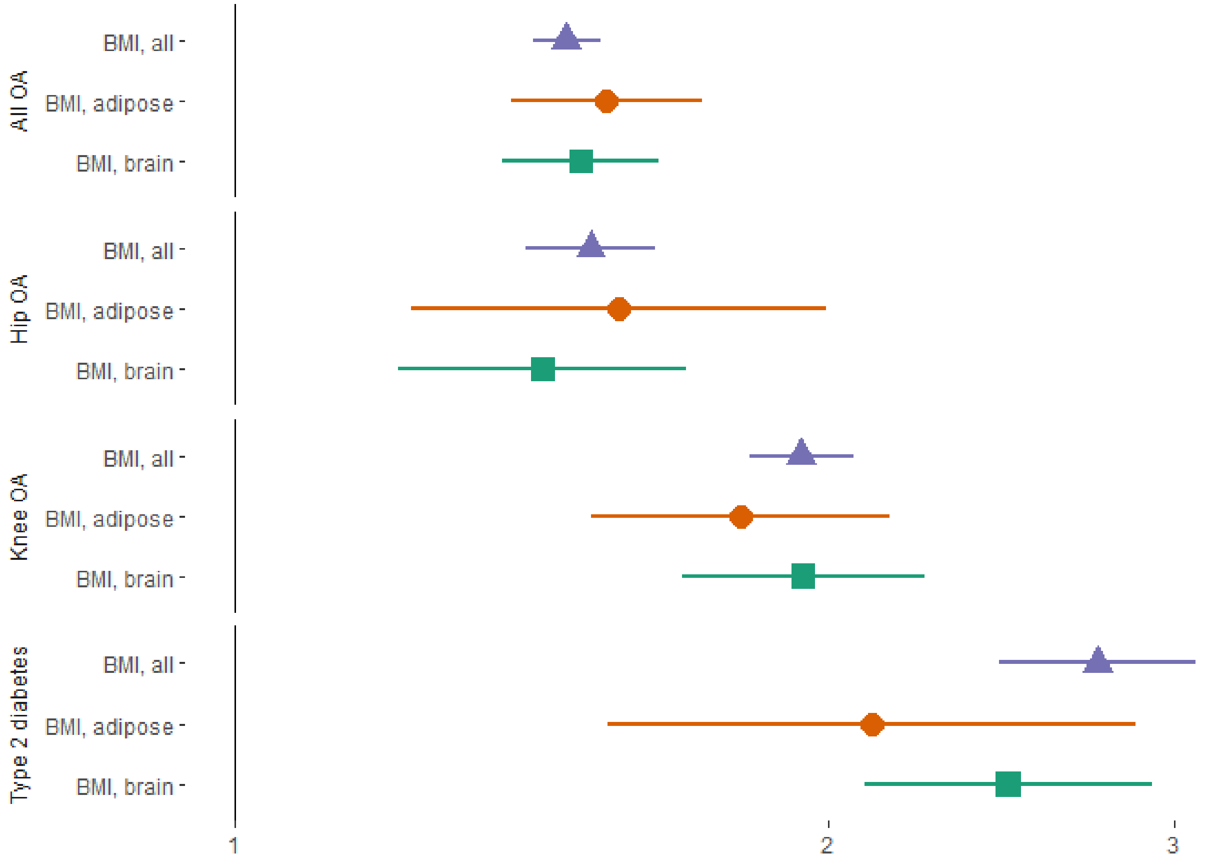


**Figure S23: Results of causal inference analyses with tissue-specific BMI instruments.** Instruments for the Mendelian randomization analysis were defined as BMI-associated variants that colocalize with eQTLs from the brain and subcutaneous adipose tissue, respectively. The y-axis depicts the magnitude of effect (odds ratio) along with the 95% confidence interval.
